# Frailty, initial attrition and feasibility of novel platinum-free options for advanced non-small-cell lung cancer in the real-world setting

**DOI:** 10.64898/2026.02.02.26345340

**Authors:** Petros Christopoulos, Miriam Blasi, Sandra Langer, Shuo Shi, Jelena Cvetkovic, Farastuk Bozorgmehr, Michael Allgäuer, Kadriya Yuskaeva, Marc A. Schneider, Rajiv Shah, Jonas Kuon, Albrecht Stenzinger, Thomas Glück, Michael Thomas

## Abstract

**Background:** How frailty limits initial therapy in metastatic non-small-cell lung cancer (mNSCLC) remains poorly understood and was investigated in this study.

**Methods:** We retrospectively analyzed 2592 consecutive patients with mNSCLC treated between 2018-2023.

**Results:** Systemic therapy was initiated in 74% of patients with PD-L1 0-49% (n=1306) *vs.* 79% with PD-L1 ≥50% (n=507, p=0.014), in whom availability of monoimmunotherapy reduced best supportive care (16.4% *vs.* 22.3%, p=0.0002), while early death remained unchanged (ca. 4%). 70% of patients receiving mBSC were initially treatable but suffered early deterioration associated with comorbidities, metastatic burden, or protracted workup (p<0.001). The atezolizumab Summary of Product Characteristics (SmPC) criteria, *i.e.* >80 years, or ECOG performance status (PS) ≥3, or comorbidities with PS ≥2 or age ≥70, were fulfilled by 38% (n=501) and associated with 3-fold higher risk of death without therapy (230/501) compared to non-SmPC patients (p<0.001). Under platinum, SmPC patients showed higher toxicity and shorter survival than non-SmPC patients, which for a platinum dose ratio ≤60% across 4 cycles (9% of 1306) resembled that with single-agent chemotherapy (median 5.1 months). SmPC criteria correlated with platinum use stronger than comorbidity scores, but predictability for individual patients remained modest (AUC 0.71, p<0.001).

**Conclusions:** The high pretherapeutic attrition of 26% in mNSCLC improved with availability of monoimmunotherapy, but requires more efficient, faster patient workflows for further mitigation. Adoption of the SmPC criteria could support identification of patients at higher risk for mBSC or potential platinum overtreatment to enhance utilization of novel platinum-free first-line options.

**Highlights:**

- 25% of NSCLC patients die without therapy mainly due to early deterioration
- SmPC criteria indicate 3x higher risk for BSC in metastatic NSCLC
- Higher toxicity and shorter survival for SmPC *vs.* non-SmPC patients under platinum
- Novel drugs and optimized workflows could cut pretherapeutic attrition to 10%
- Improved therapeutic allocation is key for upfront use of platinum-free options

## Introduction

Non-small-cell lung cancer (NSCLC) is a leading cause of cancer-related mortality globally.^1^ The prognosis is further aggravated by the advanced age and poor clinical condition of many patients at diagnosis, which limit therapeutic options, in particular the applicability of platinum-based chemotherapy.^2,3^ This poses a major therapeutic challenge, meanwhile also recognized in contemporary guidelines, because platinum still remains a key part of first-line treatment for most cases besides immune-checkpoint (ICI) and tyrosine kinase inhibitors (TKI).^4–6^ While vinorelbine monotherapy could prolong median overall survival (mOS) *vs.* best supportive care (BSC) for >70 years old NSCLC patients with Eastern Cooperative Oncology Group performance status (ECOG PS) 0-2, the benefit was small (28 *vs.* 21 weeks) and accompanied by drug-related toxicity.^7^ Moreover, platinum-free combination chemotherapies, like the administration of vinorelbine together with gemcitabine, have yielded contradictory results.^8^ On the other hand, elderly patients with a better ECOG PS 0-1 and ability to receive platinum doublets have consistently enjoyed a longer mOS compared to single-agent vinorelbine, gemcitabine or pemetrexed, *i.e.* 10.3 *vs.* 6.2, or 9.6 *vs.* 7.5 months in two exemplary studies.^9,10^ These results highlight the significant unmet need for NSCLC patients assigned to monotherapies.

Atezolizumab emerged recently as a more attractive alternative after demonstrating longer mOS (10.3 *vs.* 9.2 months) and fewer grade 3-4 related adverse events (TRAEs 16% vs. 33%) compared to single-agent vinorelbine or gemcitabine in the phase 3 IPSOS study.^11^ Consequently, the European Medicines Agency (EMA) approved atezolizumab monotherapy regardless of the PD-L1 tumor proportion score (TPS), and recommended to consider this option for potentially platinum ineligible patients based on the criteria defined in the atezolizumab Summary of medicinal Product Characteristics (SmPC): ECOG PS ≥3; or age >80 years; or ECOG PS2 / age ≥70 years in combination with comorbidities of certain system-organ classes (SOC: pulmonary, cardiac, vascular, renal, metabolism, nutrition, nervous system and psychiatric) contraindicating platinum, as assessed by the treating physician.^11,12^ These SmPC criteria were met by most patients (89%) enrolled in the IPSOS study and associated with significantly better outcomes under atezolizumab compared to single-agent chemotherapy.^13^ However, it remains unclear how prevalent and reliable the SmPC features are in the real-world setting, and how they could be integrated into existing algorithms to guide therapeutic decisions in daily clinical practice. The current study was performed to address these key questions in an unbiased manner based on evidence about SmPC status and actual treatment in a large (> 2500) all-comer patient population representative for the German and European clinical reality.

## Patients and methods

### Study design and objectives

This single-center retrospective chart review included all patients with newly diagnosed stage IV NSCLC treated from January 2018 until December 2023 at the Thoraxklinik Heidelberg. The study period was chosen to ensure both adequate follow-up and availability of pembrolizumab monotherapy for all patients with PD-L1 TPS ≥50%, which was approved by the EMA in 2017 based on the results of Keynote-24. Study endpoints were: 1) the actual therapeutic allocation and outcomes of newly diagnosed patients with an emphasis on initial attrition (defined as death after diagnosis without any systemic treatment) and inability to receive platinum; 2) the prevalence, characteristics, treatment and outcomes of the patient subset fulfilling the SmPC criteria of atezolizumab according to the EMA label based on IPSOS^12^; and 3) the ability of various clinical parameters and scores to determine platinum eligibility and the potential use of monoimmunotherapy in the real-world setting. Scoring systems that were prospectively captured as part of the routine care, or could be retrospectively calculated were used and included the Simplified Comorbidity Score (SCS), the Charlson Comorbidity Index (CCI) and the age-adjusted CCI (ACCI).^14–16^ In addition, the patients’ comorbidities were categorized according to the type and number of affected SOC among the 7 predefined categories^12^: i) cardiac; ii) vascular; iii) nervous system; iv) psychiatric; v) renal and urinary; vi) metabolism and nutrition; vii) respiratory, thoracic and mediastinal disorders. Best supportive care (BSC) was classified as based on the patient’s wish whenever patients had actively declined the treatment recommended by their physicians, as documented in the medical records in accordance with the local regulations. In all other cases, BSC was considered to reflect a shared decision between physicians and patients, typically prompted by poor clinical condition (“medical BSC”, or mBSC). On the other hand, the category of “early death” included patients, for whom a decision to proceed with systemic treatment had been documented, but they died before the planned treatment could be initiated. Further details of Methods are given in the Supplements.^17,18^

## Results

### Patient characteristics

Overall, 2592 patients admitted for stage IV lung cancer in the Thoraxklinik Heidelberg during the defined study period 2018-2023 could be identified, among which 2031 with histologically confirmed NSCLC comprised the “overall study population” (Figure 1 and Supplementary Results). Lung cancer was diagnosed or suspected either through symptomatic workup, or as an incidental finding on imaging performed for unrelated reasons, but none of the patients had undergone lung cancer screening, which was not implemented in Germany until 2025. Among 1813 patients with available PD-L1 results, 507 (28.0%) had a high PD-L1 TPS ≥50% and similar characteristics to the overall NSCLC population (Suppl. Table S1), while 1306 (72.0%) had a PD-L1 TPS 0-49% and comprised the “main study population” with detailed characteristics listed in Table 1. These showed typical NSCLC demographics, *i.e.* a predominance of male (60.7%) ex-/current smokers (91.1%) with a median age of 67.2 years. The median follow-up was 43.1 (95% confidence interval [CI] 38.3-47.9) months, and the median OS 6.7 (95% CI 5.9-7.5) months. For patients with ECOG PS 0 at the time of treatment start and no comorbidities (n=115), the median OS was 16.5 (95% CI 12.5-20.5) months across histologies. Among patients receiving any treatment, the 5-year OS rate was 13.1% (95% CI 10.4%-15.8%).

**Figure 1.**
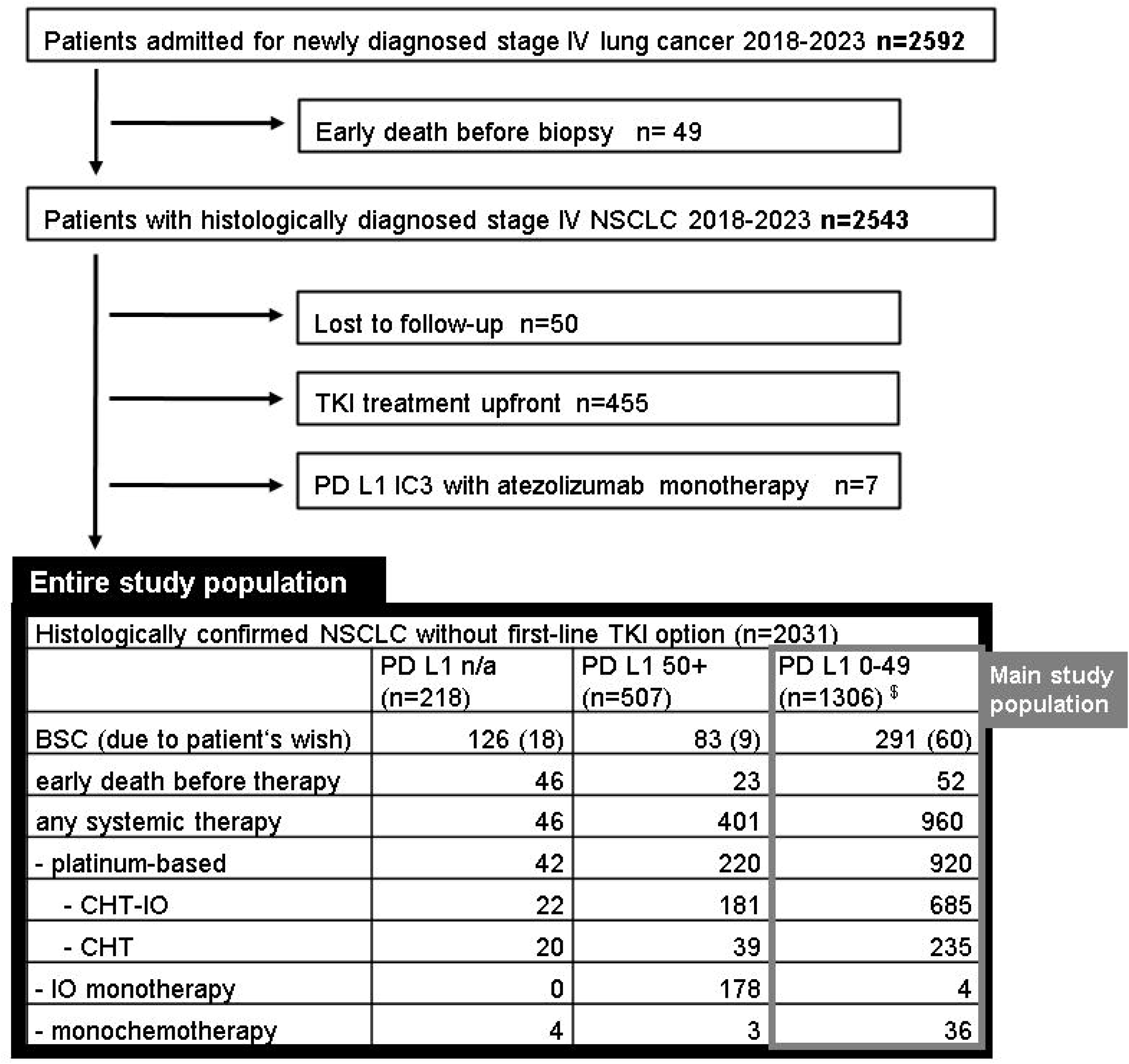
Flowchart of study patients. <u>Abbreviations:</u> BSC; Best supportive care; CHT: chemotherapy; IC3: expression on at least 10% of tumor immune cells; IO: immunotherapy; n: number; n/a: not applicable; NSCLC: non-small-cell lung cancer; PD-L1: Programmed Cell Death Ligand 1; TKI: tyrosine kinase inhibitors. ^$^: 3/1306 patients are currently in watch-and-wait for lepidic tumors with low burden after local therapy.

**Table 1.**
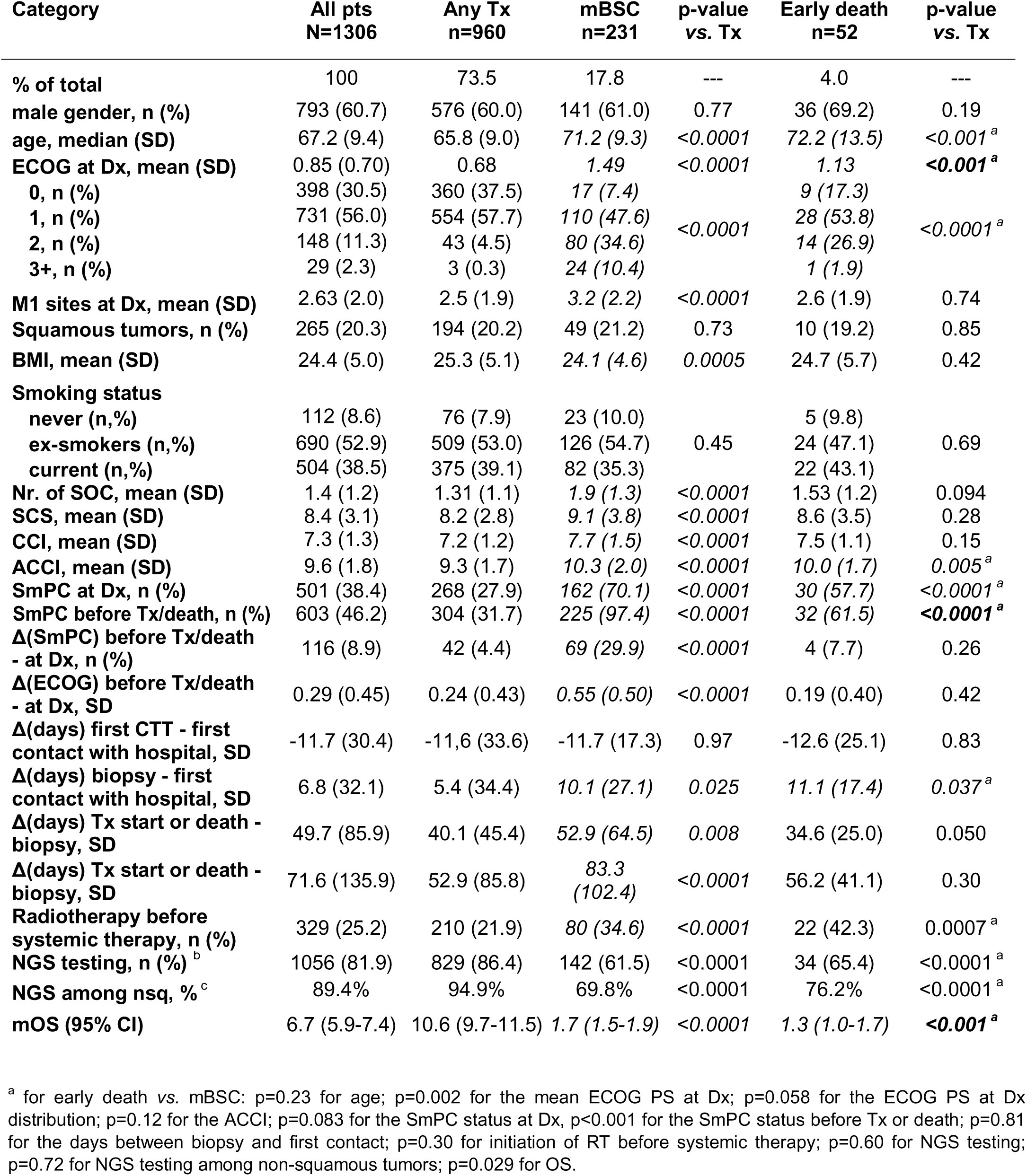
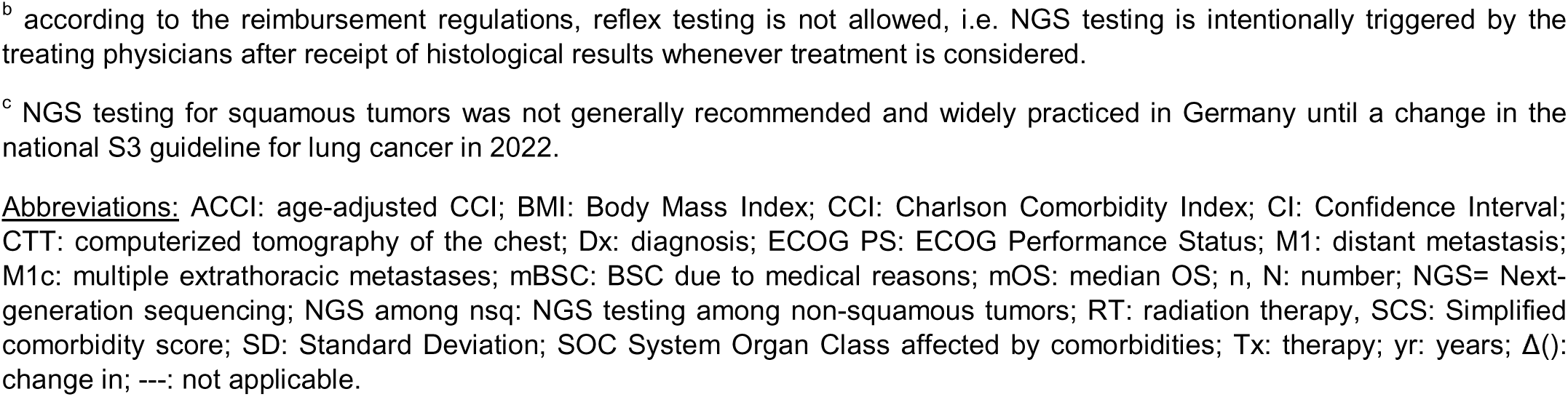
Characteristics of the main study population according to therapeutic allocation. Significant differences (p<0.05) for the comparisons “mBSC” *vs.* <u>”any Tx”,</u> and “early death” *vs.* <u>”any Tx”</u> are highlighted in italics; furthermore, for parameters with significant differences in both comparisons, those with significant difference also in the comparison “mBSC” *vs.* “early death” are highlighted in bold italics. Statistical comparisons were performed with a t-test for continuous and Chi-square test for categorical variables. A more extensive version of this Table is provided as Supplementary Table 2.

### High attrition rate before treatment start in NSCLC

Among patients with histologically confirmed NSCLC without first-line TKI, only 69.3% (1407/2031, Figure 1) were given systemic anticancer treatment. The rest received best supportive care for medical reasons (mBSC, 20% or 414/2031), or by own choice (4.2%), or suffered early death before the planned treatment could be initiated (6%). Among patients without PD-L1 testing, the pretherapeutic attrition was even higher at 79% (172/218, p<0.001) due to an increase of both BSC (58%, p<0.001) and early death (21%, p<0.001, Figure 1). An additional subset of patients admitted for suspected lung cancer had died earlier, before a biopsy could be performed (49/2592 or 1.9%, Figure 1).

### Treatment-related barriers to systemic therapy in NSCLC

Significantly more patients with PD-L1 TPS ≥50% and available monoimmunotherapy received systemic anticancer therapy compared to patients with PD-L1 TPS 0-49%, but the difference was modest at 5.4% (79.1% *vs.* 73.6%, p=0.014). BSC was significantly reduced (16.4% *vs.* 22.3%, p=0.0002), while early death remained unchanged (4.5% *vs.* 4.0%, p=0.5944, Figure 2). Interestingly, BSC due to patients’ wish decreased more (1.8% *vs.* 4.5%, *i.e.* by 60%, p=0.0049) than mBSC (14.6.% *vs.* 17.8%, *i.e.* by 18%, p=0.1060, Figure 2). Also, the type of administered treatment shifted in patients with PD-L1 TPS ≥50%, with significantly more PD-(L)1 inhibitor monotherapies (35.1% *vs.* 0.3%, p<0.0001), but fewer platinum-based regimens (43.3% *vs.* 70.4%, p<0.0001) and monochemotherapies (0.6% *vs.* 2.7%, p=0.0044, Figure 2).

**Figure 2.**
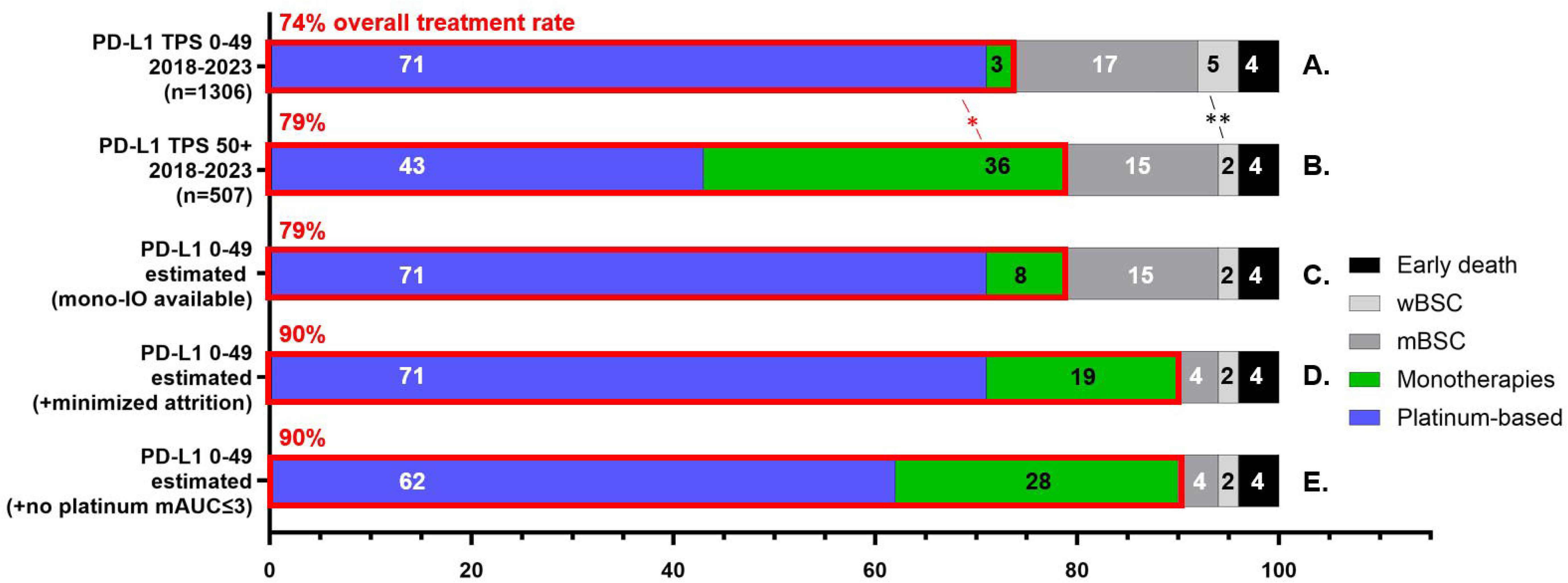
Therapeutic allocation of newly diagnosed NSCLC patients. Empiric data for patients with PD-L1 TPS 0-49% before approval of monoimmunotherapy (**A**), and for patients with PD-L1 TPS ≥50% after approval of pembrolizumab monotherapy (**B**) are based on Figure 1. The expected allocation for patients with PD-L1 TPS 0-49% and availability of monoimmunotherapy, but the same platinum utilization and no additional measures against initial attrition (**C**); with availability of monoimmunotherapy and measures to enable therapy for the 70% of mBSC patients who are treatable early after diagnosis based on manual ordering of NGS and PD-L1 by the physicians (**D**), as well as additional avoidance of platinum in SmPC patients who achieve only a very low average carboplatinum AUC ≤3 dose with excessive toxicity and poor survival (**E**) are estimates based on findings of this study. Further details are given in the Results and Discussion. The numbers in Figure 2 have been rounded. *: p<0.05; **: p<0.01 with a Chi-square test. <u>Abbreviations:</u> mAUC: carboplatinum average area under the curve across 4 cycles; IO: immunotherapy, mBSC: best supportive care due to medical reasons; PD-L1 TPS: Programmed Cell Death Ligand 1 Tumor Proportion Score; SmPC: Summary of medicinal Product Characteristics for atezolizumab; wBSC: best supportive care due to the patient’s wish.

### Patient-related and systemic barriers to anticancer therapy in NSCLC

As mBSC (*ca.* 15%) and early death (*ca.* 4%) were not significantly affected by the availability of monoimmunotherapy, we explored treatment-unrelated obstacles therapy initiation in NSCLC. More advanced age >70 years (p<0.001), a worse ECOG PS (p<0.0001), more comorbidities (evident as higher ACCI/CCI/SCS, p<0.001), and fulfillment of the SmPC criteria (“SmPC cases”, p<0.0001) were uncontrollable patient-related factors associated with initial attrition (Table 1 and Figure 3). Furthermore, while all patients had a first contact with our hospital approximately 11-12 days on average after initial chest CT with suspicion of lung cancer, those with subsequent mBSC or early death had a longer time from first contact to biopsy (+5 days on average, p<0.05), as well as a more frequent initiation of radiotherapy before systemic therapy (+15 or +20%, p<0.001) compared to patients who subsequently received systemic treatment (Table 1 and Figure 3D). During this time, many patients with subsequent mBSC deteriorated, as evident by a significant increase in the ECOG PS from initial contact until before treatment start or death (Table 1, Suppl. Figure S1, Suppl. Table S3): for example, 80 non-SmPC patients (6.1% of 1306 main study patients, or 10% of 805 non-SmPC patients) could not receive platinum-based therapy due to early clinical deterioration caused, *e.g.*, by tumor progression or infection (Suppl. Table S4). On the other hand, patients with subsequent early death had started with a better ECOG PS than mBSC patients, but had shorter survival due to acute complications, e.g. sepsis or pulmonary embolism, without the evidence of progressive deterioration, *i.e.* change in ECOG PS or SmPC status, noted for mBSC patients (Table 1). For most patients with mBSC or early death, the initial intention to administer systemic therapy was evident by the manual triggering of NGS testing by the treating physician in ≥60% of all and ≥70% of non-squamous tumors (Table 1).

**Figure 3.**
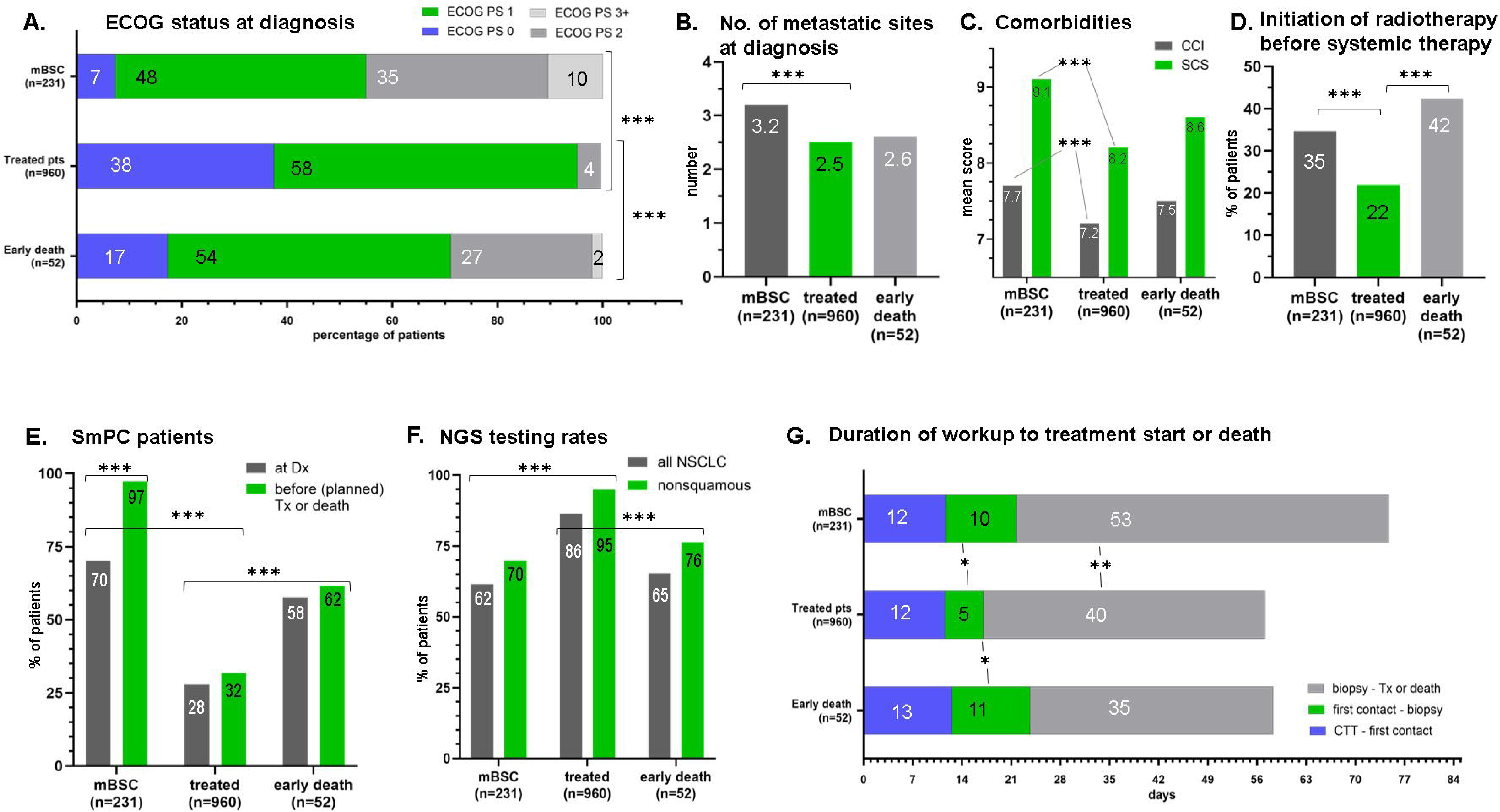
Key factors associated with mBSC and early death among NSCLC patients with PD-L1 TPS 0-49%. (**A**) Distribution of ECOG PS at initial diagnosis compared to patients that subsequently received systemic anticancer treatment; (**B**) The number of metastatic sites at initial diagnosis; (**C**) The average Charlson Comorbidity Index and Simplified Comorbidity Score at initial diagnosis; (**D**) The percentage of patients with initiation of radiotherapy before systemic therapy; (**E**) The percentage of patients fulfilling the SmPC criteria at the time of initial diagnosis *vs.* before (planned) first-line treatment or death; (**F**) NGS testing rates among patients with tumor of any histology or non-squamous tumors (NB. NGS testing of squamous tumors was not generally recommended in Germany until 2022); (**G**) Duration of various phases in the workup of patients until initiation of first-line treatment or death. *: p<0.05; **: p<0.01; ***: p<0.001 with a Chi-square test for categorical, and t-test for continuous variables. Further details, including confidence intervals, are given in Table 1 and Suppl. Table S2. <u>Abbreviations:</u> CTT: computerized tomography of the chest (thorax); CCI: Charlson Comorbidity Index; Dx: diagnosis; ECOG PS: ECOG performance status; mBSC: Best supportive care due to medical reasons; NGS: next-generation sequencing; NSCLC: non-small-cell lung cancer, PD-L1 TPS: Programmed Cell Death Ligand 1 Tumor Proportion Score; SCS: Simplified Comorbidity Score; SmPC: Summary of medicinal Product Characteristics for atezolizumab. Tx: therapy.

### SmPC patients in the real-world setting

SmPC patients comprised 38.4% (501/1306) of the main study population and showed a very low treatment rate of 53.5%, with significant overrepresentation of all 3 categories of attrition, *i.e.* early death (relative risk [RR] 2.3), mBSC (RR 4.0) and BSC by patients’ choice (RR 2.8, Figure 4A and Suppl. Table S5). As monotherapies were rare, almost 90% of treated SmPC patients received platinum (238/268), but showed significantly shorter OS (9.6 *vs.* 11.3 months in median, p=0.012) and higher toxicity than platinum-treated non-SmPC patients: fewer platinum cycles, a lower cumulative dose ratio, more platinum dose reductions, more interruptions, and more discontinuations (all p<0.001, Figure 4B and Suppl. Table S6), which were all significantly associated with shorter OS among platinum-treated patients (Figure 4C). Among those characteristics, the average platinum dose ratio across the 4 chemotherapy cycles showed the strongest association with shorter OS of platinum-treated SmPC patients (HR 5.70; p<0.001, Figure 4C). SmPC patients with platinum dose ratio above the median of 61.5% (corresponding to carboplatinum AUC >3) showed significantly longer mOS (15.5 months), similar to that of platinum-treated patients, while the shorter OS of SmPC patients with a lower platinum dose (5.1 months, p<0.001) was similar to that of patients receiving monotherapies (5.1 months, Figure 4D). Furthermore, SmPC patients with a lower platinum dose showed more dose reductions of the first cycle (74% *vs.* 51%, p <0.001), more dose reductions and (24% *vs.* 3%, p<0.001) or interruptions/discontinuations (98% *vs.* 82%, p<0.001) during the entire platinum course, more platinum discontinuations before the 4^th^ cycle due to toxicity followed by other treatment (45% *vs.* 8%, p <0.001) or death (27% *vs.* 1%, p <0.001), and more discontinuations due to radiologic tumor progression (13% *vs.* 3%, p<0.0001) than SmPC patients with a higher platinum dose, whose treatment characteristics were comparable to those of platinum-treated non-SmPC patients (Figures 4E, 4F and Suppl. Table S7). Further details are given in the Supplementary Results.^17,18^

**Figure 4.**
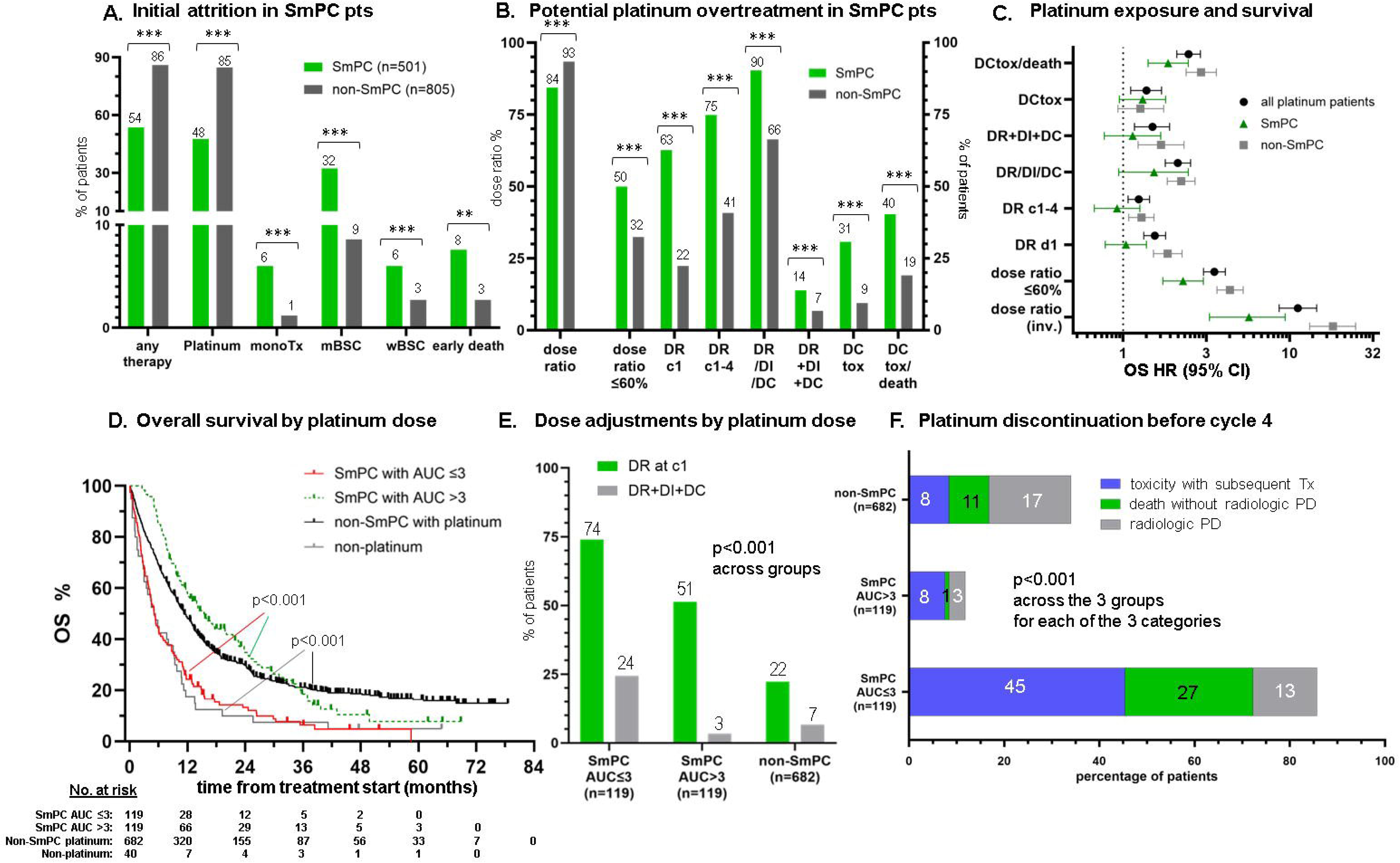
Management, toxicity and survival of SmPC patients in the real-world setting. (**A**) Overall rate of treatment and use of various therapies in SmPC vs. non-SmPC patients (details in Suppl. Table S5); for SmPC patients the relative risk (RR) of dying without systemic treatment was 3.31 (95% CI 2.54-4.32), the RR of not receiving platinum was 3.43 (95% CI 2.65-4.46), the RR of mBSC was 4.00 (95% CI 2.92-5.46), the RR of early death was 2.32 (95% CI 1.32-4.07) and the RR for BSC by patients’ wish was 2.80 (95% CI 1.63-4.78) compared to non-SmPC patients. (**B**) Comparison between platinum-treated SmPC and non-SmPC patients regarding: platinum dose ratio across the planned 4 cycles (as % of the full dose); percentage of patients with platinum dose ratio ≤ 60%; percentage of patients with platinum dose reduction initially (at c1) or at any time during the 4 cycles (C1-4); percentage of patients with platinum dose reduction or interruption or discontinuation during the 4 chemotherapy cycles; percentage of patients with platinum dose reduction and interruption and discontinuation during the 4 chemotherapy cycles; percentage of patients with platinum discontinuation due to toxicity before cycle 4 with subsequent further treatment; percentage of patients with platinum discontinuation due to toxicity before cycle 4 with subsequent further treatment or death. (**C**) The association of platinum dose ratio (inverse), platinum dose adjustments and platinum discontinuation with overall survival of platinum-treated patients; (**D**) Overall survival of platinum-treated SmPC patients by platinum dose compared to platinum-treated non-SmPC patients and patients receiving platinum-free therapies (mostly monotherapies, details are given in Suppl. Table S8): median OS was 5.1 (95% CI 3.9-6.3) months for SmPC patients with average AUC ≤3 (n=119), 15.5 (11.5 – 19.5) months for SmPC patients with average AUC >3 (n=119), 11.3 (10.1 – 12.5) months for platinum-treated non-SmPC patients (n=682) and 5.1 (2.9 – 7.3) for patients who received monotherapies (log-rank p<0.001); (**E**) Frequency of platinum dose adjustments for SmPC patients who received a low average carboplatinum AUC ≤3 dose across the four cycles, *vs.* SmPC patients receiving higher average platinum doses, *vs.* platinum-treated non-SmPC patients; (**F**) Overall frequency and reasons of platinum discontinuation before the 4^th^ cycle by patient subgroup. Further details are given in Suppl. Tables S5 and S6; *: p<0.05; **: p<0.01; ***: p<0.001 with a Chi-square for categorical, t-test for continuous, and Cox regression (4C) or logrank test (4D) for survival data. <u>Abbreviations:</u> AUC: area under the curve for carboplatin dose; BSC: best supportive care (m: due to medical reasons; w: due to patients’ wish); c1-4: cycles 1-4; CI: confidence interval; DC: platinum discontinuation; DCtox: platinum discontinuation due to toxicity before cycle 4 with subsequent further treatment; DCtox/death: platinum discontinuation before cycle 4 with subsequent further treatment or death; :DI: dose interruption; DR: dose reduction; HR: hazard ratio; OS: overall survival; SmPC: Summary of Product Characteristics for atezolizumab; Tx: therapy.

### Predictability of upfront monoimmunotherapy for NSCLC with PD-L1 TPS 0-49%

For the identification of NSCLC patients foregoing platinum and thus potentially suitable for monoimmunotherapy, the SmPC criteria at diagnosis, as a composite characteristic, outperformed individual parameters, such as age, ECOG PS, various comorbidities scores, and other clinical and laboratory parameters (Suppl. Table S9). However, their discriminatory performance for individual patients remained moderate with a sensitivity of 64%-68% and a specificity of 74%-76% when used alone (AUC 0.71, Gini index 0.42), or in conjunction with available NGS results among all patients (AUC 0.70, Gini index 0.40), or for non-squamous tumors (AUC 0.70, Gini index 0.39, Suppl. Table S9).

Among platinum-treated patients, the SmPC criteria were significantly linked to platinum discontinuation (Figure 4) with modest predictive performance for individual patients (sensitivity 43%-47%, specificity 77-80%, AUC 0.61, Gini index 0.23), which was, however, not worse than that of the most informative other clinical variables, like the ACCI score (Suppl. Table S10). Among platinum-treated SmPC patients, no clinical parameter retained noteworthy predictive capacity, either for platinum discontinuation (AUC 0.41-0.57, Suppl. Table S10), or for a low average carboplatinum dose AUC ≤3 as surrogates of potential platinum overtreatment (AUC 0.41-0.55, Suppl. Table S11).

## Discussion

One main finding of the current study was the high attrition rate of approximately 25% for NSCLC patients already before the start of systemic therapy. While a similar attrition rate of ca. 25-35% has been described between subsequent therapy lines for EGFR+/ALK+ or wild-type NSCLC,^19–21^ early patient losses before the first line have not been a focus of research yet. Systematic studies are lacking, while occasional estimates in the literature are scarce and partly contradictory, for example 59% in a very old study conducted >20 years ago *vs.* 27% in a recent series from Canada which, however, described radiotherapy as the main first-line option for 44% of stage IV NSCLC.^22,23^ According to our results, this initial attrition is mainly caused by frailty and/or aggressive disease, which facilitate rapid clinical deterioration during prolonged workup or upfront radiotherapy, so that initial plans for systemic treatment are canceled by early death or replaced by mBSC. Related clinical characteristics included an older age, worse ECOG PS, more comorbidities, and more advanced metastatic spread at diagnosis (Table 1). This is remarkable for NSCLC, which is generally regarded as a relatively slow-growing tumor and has not traditionally carried the same sense of urgency to initiate treatment as soon as possible that we associate, for example, with SCLC or acute leukemia.^24,25^ The current results, however, clearly show that devastating complications and deterioration precluding treatment can occur very early in NSCLC, despite the slower biologic tumor growth. Therefore, our timeline requirements for initial management warrant reconsideration, especially as novel immunotherapeutic and targeted drugs can now facilitate long-term survival for many patients.^26,27^

The exact type of planned therapy appears to influence attrition, but to a limited extent. With availability of monoimmunotherapy, early patient losses before treatment were reduced by a modest 5% for NSCLC with PD-L1 TPS ≥50% (Figure 2). BSC due to the patients’ wish was reduced more substantially (by 60%) than mBSC (by 18%), which is plausible given that qualitative studies have identified anticipated treatment-related suffering and toxicity as key reasons for the refusal of cancer treatment,^28^ and have also demonstrated a greater preference among patients with lung cancer for immunotherapy *vs.* chemotherapy.^29^ Consistent with this notion, attrition between therapy lines in ALK+ NSCLC has been reported lower before next-line TKI (ca. 25%) than before next-line chemotherapy (ca. 33%).^20^ Hence, availability of atezolizumab as a platinum-free option for NSCLC with PD-L1 TPS 0-49% could help more elderly and frail patients receive systemic treatment by increasing both their acceptance for the proposed anticancer therapies and their suitability for accessible options. However, even with availability of monoimmunotherapy for NSCLC with TPS ≥50%, the pretherapeutic attrition remains high > 20% (Figure 2).

Optimized, faster workflows will be essential to mitigate early deterioration after diagnosis as a key factor that precludes therapy in many patients. Delays until biopsy (Figure 3) could be mitigated by earlier appointments for the necessary radiologic examinations and bronchoscopy, which, however, would require costly increases in the capacity of the respective facilities. Beyond structural changes, a simpler measure would be adoption of liquid biopsies for initial genetic profiling, as these eliminate the waiting time for invasive tissue sampling procedures and have already demonstrated reliability and clinical utility in several prospective trials, but are not reimbursed in many countries yet.^30–32^ Delays after biopsy could be mitigated by shortening the NGS turnaround time, for example by using fully automated instruments,^33^ expanding capacity and improving patient allocation across clinics and hospitals to expedite appointments for treatment initiation, and deferring radiotherapy until afterward. All these measures together could potentially cut pretherapeutic attrition for metastatic NSCLC down to ca. 10% by enabling treatment for approximately 70% of patients who currently receive mBSC, but were treatable early after diagnosis based on manual ordering of NGS by the treating physicians (Figure 2D and Table 1). To what extent an earlier treatment start could improve survival remains to be seen. However, reducing initial attrition will become increasingly important in the near future, as novel, more potent platinum-free options, like bispecific antibodies and antibody-drug conjugates, become available for first-line use and demonstrate the ability to control aggressive tumors, which may be refractory to currently available options.

Furthermore, the current study highlights the potential clinical utility of SmPC criteria as a simple, practical tool to improve therapeutic allocation in NSCLC. Ascertaining the SmPC status on a newly diagnosed NSCLC patient represents a red flag of 3x higher risk for mBSC that should trigger swift workup and initiation of systemic therapy as soon as possible, as almost half of them (46.5%) currently die without systemic therapy (Figure 4A). On the other hand, roughly the other half (47.5%) of SmPC patients receive platinum, but most (>2/3) will discontinue before the 4^th^ cycle due to toxicity or death (Figure 4F). Among platinum-treated SmPC patients, 50% will be able to receive only a low average carboplatinum dose AUC ≤3 across the 4 cycles, with increased toxicity and poor outcomes comparable to these of patients receiving single-agent chemotherapy (Figure 4 and Suppl. Table S7). This administration of platinum for SmPC patients with excessive toxicity, but poor survival can be considered as potential platinum overtreatment and affects approximately 9% of all patients with PD-L1 TPS 0-49% (Figure 2E). Monoimmunotherapy would arguably be a preferrable option in this setting, as atezolizumab showed better efficacy and tolerability than monochemotherapies for SmPC patients in the IPSOS trial.^11^ While it remains unclear whether immunotherapy could prolong survival *vs.* dose-reduced platinum in these unfavorable cases, it would at least be less toxic. Options include atezolizumab, as approved by the EMA regardless of PD-L1 expression based on IPSOS, or pembrolizumab, as approved by the FDA based on the Keynote-42 trial for PD-L1 TPS 1-49, but not for PD-L1 negative NSCLC. Besides PD-L1 expression, which is an established biomarker of immunotherapeutic efficacy,^34,35^ predictive biomarkers of platinum toxicity could also be used to guide stratification of platinum-eligible SmPC patients, but remain an unmet need, as clinical parameters lose their predictive capacity after SmPC preselection, according to our results (Suppl. Tables 10 and 11). Comprehensive geriatric assessment (CGA) and prospective scores, like CARG and CRASH, could assist here: ^36–38^ SmPC patients presenting as platinum candidates with low scores (*e.g.* CARG ≤5) could presumably proceed safely to platinum, for those with high scores (*e.g.* CARG ≥10) caution and discussion of monoimmunotherapy is warranted, while in case of intermediate scores the decision could be individualized according to the patients’ preference. While ICI can cause immune-related adverse events in approximately 30% of patients with an equal frequency between men and women,^39,40^ as well as infusion-related reactions, hyponatremia and inappetence which may further aggravate the already impaired prognosis of many frail patients with malnutrition according to some recent metaanalyses,^41–44^ chemotherapy is generally more problematic. Consistent with this notion, prospective CGA of newly diagnosed NSCLC patients in a previous study was associated with a lower rate of platinum use (46%) and at the same time also with a lower rate of BSC (23%) compared to our real-world patients.^45^ Further criteria to stratify platinum-eligible SmPC patients in the future could include single-nucleotide polymorphisms associated with platinum toxicity, or serum levels of platinum after a short initial therapeutic trial.^46,47^

The main limitation of the current study is its retrospective character, which cannot exclude confounding and could not capture activities of daily living and other necessary elements of various chemotoxicity scores. Furthermore, as none of our patients was diagnosed in the context of lung cancer screening, it remains unclear whether the high attrition observed in our study would also affect patients identified at the asymptomatic stage through the low-dose CT programs that now roll out in several European countries based on the NELSON results.^48^ Specific strengths are the very large number of >2500 patients homogenously treated in the same institution representative for German and European practice, state-of-the-art molecular diagnostics, and deep clinical annotation of baseline characteristics, comorbidities, treatment details and outcomes to analyze the real-world characteristics of SmPC patients and address the problem of therapeutic allocation for NSCLC in a broader manner than previous studies.

In summary, we observed a high initial attrition of approximately 25% among NSCLC patients after diagnosis, which is mainly caused by clinical deterioration before treatment and would only modestly (by ca. 5%) improve through the mere availability of monoimmunotherapy. Optimized, faster patient workflows, wider adoption of liquid biopsies and long-term structural improvements in the health care system will be essential for proper containment. The integration of SmPC criteria in daily algorithms could support *a priori* identification of newly diagnosed patients at higher risk for potential platinum overtreatment (ca. 10%) or death without systemic therapy (ca. 20%). Further dissection of platinum ineligibility and improvement of initial therapeutic allocation in NSCLC will acquire greater importance as additional novel platinum-free options, like bispecific antibodies and antibody-drug conjugates become available for first-line use in the future.

## Clinical Practice Points

- There is a high pretherapeutic attrition of approximately 25% in advanced NSCLC, which is mainly caused by rapid clinical deterioration early after diagnosis and resembles the well-described later attrition between successive treatment lines.
- The rate of treatment increases only modestly, by approximately 5%, by the mere availability of monoimmunotherapy, and more efficient, faster patient workflows will be needed for further improvements.
- The SmPC criteria in newly diagnosed patients with advanced NSCLC indicate a higher risk for death without systemic treatment, as well as for increased toxicity and shorter survival under platinum, compared to non-SmPC patients.
- Measures to contain initial attrition and improve therapeutic allocation will be essential for the utilization of any novel platinum-free first-line option in the future.

## Data Availability

All data produced in the present study are available upon reasonable request to the authors

## Acknowledgements

This study was funded by the German Center for Lung Research (DZL) and Roche. Medical writing assistance was provided by Dr. Kirsten Dahm, AMS Advanced Medical Services GmbH. Mannheim, Germany.

## Authors’ disclosures of potential conflicts of interest

PC: research funding from AstraZeneca, Amgen, Merck, Novartis, PharmaMar, Roche, and Takeda; speaker’s honoraria from AstraZeneca, Gilead, Johnson & Johnson, Merck, Novartis, Pfizer, Roche, Takeda, and ThermoFisher; support for attending meetings from AstraZeneca, Daiichi Sankyo, Eli Lilly, Gilead, Johnson&Johnson, Merck, Novartis, Pfizer, and Takeda; and personal fees for participating in advisory boards from AstraZeneca, Boehringer Ingelheim, Chugai, MSD, Novartis, Johnson & Johnson, Pfizer, Roche, and Takeda; all outside the submitted work.

MB: speaker’s honoraria from Roche

SL: Roche employee, owner of profit participation certificates

SS: travel support from Accord Healthcare

MA: compensation for advisory board AbbVie; speaker’s honoraria from Boehringer Ingelheim

MAS: institutional funding from GSK and the German Federal Ministry of Research, Technology and Space (BMFTR).

RS: speaker honoraria from AstraZeneca, Bristol Myers Squibb, Johnson & Johnson, Neoconnect, Roche, Sanofi Aventis

JK: consultations fees from Pfizer, Roche, AstraZenca, BeOne, Bristol Myers Squibb; research support from Bristol Myers Squibb, AstraZeneca.

AS: advisory Board/speaker’s honoraria from Agilent, Aignostics, Amgen, Astellas, AstraZeneca, Bayer, Bristol Myers Squibb, Eli Lilly, Illumina, Incyte, Janssen, MSD, Novartis, Pfizer, Qlucore, QuiP, Roche, Sanofi, Seagen, Servier, Takeda, ThermoFisher; research grants from Bayer, Bristol Myers Squibb, Chugai, Incyte, MSD.

TG: Roche employee, owner of profit participation certificates.

MT: consultancy fees from Amgen, AstraZeneca, BeOne, Bristol-Myers Squibb, Boehringer Ingelheim, Daiichi Sankyo, Gilead, GlaxoSmithKline, Johnson&Johnson, Lilly, Merck, MSD, Novartis, Pfizer, PharmaMar, Pierre Fabre, Regeneron, Roche, Sanofi, Takeda; research grants from AstraZeneca, Bristol Myers Squibb, Johnson&Johnson, Merck, PharmaMar, Roche, Takeda; non-financial support (travel costs) from AstraZeneca, Bristol Myers Squibb, Boehringer Ingelheim, Daiichi Sankyo, GlaxoSmithKline, Johnson&Johnson, Lilly, Merck, MSD, Novartis, Pfizer, Pierre Fabre, Regeneron, Roche, Sanofi, Takeda.

All remaining authors have declared no conflicts of interest.

