## Supplementary Material for "Frailty, initial attrition and feasibility of novel platinum-free options for advanced non-small-cell lung cancer in the real-world setting"

### Supplementary Methods

Histopathological diagnosis was based on standard WHO criteria. PD-L1 TPS was assessed by immunohistochemistry on a Ventana system using the clone SP263, while testing for actionable genetic alterations in *EGFR, ALK, ROS1, RET, NTRK, BRAF, KRAS, HER2,* and *MET* was performed by combined DNA and RNA next-generation sequencing (NGS) using a 40-gene panel as previously published [17] (NB. NGS testing was not generally recommended for squamous tumors before 2022 in Germany). According to the local reimbursement regulations, reflex testing is not allowed, *i.e.* PD-L1 and NGS testing need to be intentionally triggered by the treating physicians after receipt of histological results whenever treatment is considered. Patient data, including demographic characteristics, weight, height, baseline ECOG PS, smoking status, comorbidities, tumor histology and metastatic locations, details of systemic anticancer treatment, various laboratory test results, as well as the dates of initial diagnosis, treatment start, tumor progression, and last follow-up or death were collected from the patients' records, with a data cut-off on January 31, 2025. The time of the first computerized tomography (CT) indicating lung cancer, which was usually performed externally before referral to our institution, was chosen as the first milestone in the patients' trajectory, as lung cancer was often an incidental finding on imaging studies performed for unrelated reasons without a previous initial visit to a primary care physician. Disease progression was determined by regular imaging via chest/abdominal CT and brain MRI every 6-12 weeks according to the standard of care. Tumor responses and the date of tumor progression according to RECIST v1.1 were verified through evaluation of radiologic images by the investigators [18]. For treated patients, progression-free survival (PFS) and overall survival (OS) were analyzed from the date of treatment start according to Kaplan-Meier, while for patients who died without therapy, OS was calculated from the time of initial diagnosis. For patients with platinum discontinuation, discontinuations due to side effects followed by further treatment were considered separately from discontinuations followed by death, as it is impossible to distinguish treatment toxicity from tumor progression *vs.* complications as the cause of death in such cases. This study was approved by the ethics committee of the Medical Faculty of the University of Heidelberg (S-659/2024), while all patients gave written consent for the collection and scientific analysis of their clinical data.

Descriptive statistics included mean or median values with standard deviations or standard errors or interquartile ranges, as appropriate. Chi-square and t-tests were used for the comparison of categorical and continuous data, respectively, while receiver operating characteristic (ROC) curves with calculation of the area-under-the-curve (AUC) and Gini index were used to assess the predictive performance of various parameters. The effect of categorical and continuous variables on survival was analyzed according to Kaplan Meier with log-rank tests and Cox regression. Duration of follow-up was calculated using the reverse Kaplan-Meier method. All confidence intervals and p-values are presented to highlight aspects of the observed data and should be interpreted in an exploratory context. Statistical calculations were performed with SPSS v30 (IBM, Armonk, NY, USA) and the programming language R version 4.1.2. Plots were generated with R v4.1.2, SPSS v30 and GraphPad Prism version 8 (La Jolla, CA, USA).

### Supplementary Results

#### Study population

Overall, 2592 patients admitted for stage IV lung cancer in the Thoraxklinik Heidelberg during the defined study period 2018-2023 could be identified. Patients who died without biopsy and histologic confirmation (n=49), were lost-to follow-up after biopsy (n=50), received targeted therapies upfront (n=455), or received atezolizumab monotherapy due to high PD-L1 expression in immune cells (IC3) despite PD-L1 TPS <50% (n=7) were excluded from further analyses. The remaining 2031 patients with histologically confirmed NSCLC comprised the overall study population (Figure 1). Among them, the PD-L1 status was not determined in 218 (10.7%), most of whom died without any systemic therapy (172/218 or 78.9%). Most of the remaining 46 patients were empirically treated with platinum regimens (42/46), either with (n=22) or without immunotherapy (n=20, mainly due to either contraindications for immunotherapy [n=4] or large-cell neuroendocrine lung carcinoma histology [n=11] under platinum/etoposide, for which combination with immunotherapy was not yet approved), while 4 frail patients received monotherapies. The main reason for lack of PD-L1 testing was insufficient tumor tissue (n=31), combined with high procedural risk and/or urgent need for treatment start (*e.g.* due to superior vena cava syndrome), and/or a failed tissue rebiopsy. NSCLC patients without PD-L1 testing were significantly older with a significantly worse ECOG PS than patients with available PD-L1 results (Suppl. Table S1). Initial radiotherapy before systemic therapy tended to be more frequent among patients with available (38.7% or 55/142) *vs.* patients without available NGS results (28.1%, or 25/89, p=0.098).

**SmPC patients in the real-world setting**

Most prevalent SmPC characteristic was older age ≥70 years with at least 1 SOC comorbidity (in 84.0% of SmPC and 32.2% of 1306 study patients), while most relevant for the patients' survival was ECOG PS ≥2 (in 29.3% of SmPC and 13.6% of 1306 study patients), which was associated with an mOS uniquely <2 months *vs.* >3 months for all other SmPC categories (Suppl. Table S5).

Non-platinum therapies were significantly more frequent in SmPC (5.4%) compared to non-SmPC patients (1.1%, p<0.001), but overall rare and associated with significantly shorter mOS (Figure 4) and mPFS (Suppl. Figure S4) than platinum-based regimens. Further details about the non-platinum therapies of study patients are given in Suppl. Table S8.

While the SmPC criteria miss younger patients (<70 years) with special contraindications for platinum, such as chronic renal failure or severe coronary artery disease, this constellation is very rare and affected <1% of our non-SmPC cases (Suppl. Table S4). Likewise, contraindications to immunotherapy are rare, observed in 4.7% of our patients (Suppl. Figure S2), and do not substantially influence the estimates shown in Figure 2.

All SmPC patients with a low platinum dose received carboplatinum, so that the average dose ratio threshold of 61.5% across 4 cycles corresponds to carboplatinum AUC ≤3. The unfavorable group with a lower dose ratio included all 12/12 SmPC patients who started carboplatin with AUC3, most (76/137 or 55%) SmPC patients who started carboplatin with

AUC4 (*i.e.* all patients starting with AUC4 who could not complete 4 cycles of treatment), and approximately 1/3 (31/89 or 35%) of SmPC patients who started with AUC5 (*i.e.* all patients starting with AUC 5 who received only 1 or 2 treatment cycles, Suppl. Figure S3). SmPC patients with a low average carboplatinum dose AUC ≤3 had a PFS comparable to that of patients receiving monotherapies, while the the PFS of SmPC patients receiving higher platinum doses was comparable to that of platinum-treated non-SmPC patients (Suppl. Figure S4).

### Supplementary Tables

| | All NSCLC patients N=2543 | PDL-1 TPS ≥50 % w/o targeted 1L N=507 | p-value vs. all patients | PDL-1 TPS n/a N=218 | p-value vs. all patients |
|---|---|---|---|---|---|
| **n (%)** | 2543 (100) | 573 (19.9) | --- | 218 (8.6) | --- |
| **Gender, males, n (%)** | 1470 (57.8) | 306 (60.4) | 0.29 | 141 (64.7) | 0.0482 |
| **Age at diagnosis, years (median, IQR)** | 67.1 (13.3) | 66.8 (16.6) | 0.19 | 69.2 (15.6) | 0.0024 |
| **ECOG PS at initial diagnosis, mean (SD)** | 0.85 (0.70) | 0.85 (0.71) | --- | 1.56 (0.85) | --- |
| **0 (n, %)** | 783 (30.8) | 155 (30.6) | | 15 (6.9)) | |
| **1 (n, %)** | 1345 (52.9) | 283 (55.8) | | 97 (44.5) | |
| **2 (n, %)** | 313 (12.3) | 59 (11.6) | 0.311 | 78 (35.8) | < 0.00001 |
| **3 (n, %)** | 79 (3.1) | 9 (1.6) | | 24 (11.0) | |
| **4 (n, %)** | 23 (0.9) | 2 (0.4) | | 4 (1.8) | |
| **Height, mean (SD)** | 170.7 (9.2) [a] | 171.1 (9.2) [b] | 0.46 | 170.5 (9.7) [c] | 0.70 |
| **Weight, mean (SD)** | 73.7 (16.5) [a] | 74.3 (16.0) [b] | 0.49 | 74.5 (17.1) [c] | 0.52 |
| **BMI, mean (SD), kg/m²** | 25.2 (4.9) [a] | 25.3 (4.4) [b] | 0.77 | 25.5 (5.0) [c] | 0.37 |

**Supplementary Table S1. Characteristics of all patients with histologically confirmed NSCLC and the PD-L1 TPS ≥50% or PD-L1 TPS n/a subsets.**

[a] All patients: Height 29 n/a, Weight 29 n/a, BMI 37 n/a; [b] PDL-1 TPS ≥50% population: Height 2 n/a, Weight 5 n/a, BMI 5 n/a; [c] PDL-1 TPS n/a population: Height 20 n/a, Weight 20 n/a, BMI 20 n/a.

Statistical comparisons were performed with a t-test for continuous and Chi-square test for categorical variables.

<u>Abbreviations</u>: BMI: Body Mass Index; ECOG PS: ECOG Performance Status; IQR: interquartile range; N, n: Number of patients; n/a: not available; NSCLC: Non-small-cell lung cancer; PD-L1: Programmed Death-Ligand 1; TPS: Tumor Proportion Score; SD: standard deviation; w/o: without; --- : not applicable.

| Category | All pts N=1306 | Any Tx n=960 | mBSC n=231 | p-value vs. any Tx | Early death n=52 | p-value vs. any Tx |
|---|---|---|---|---|---|---|
| % of total | 100 | 73.5 | 17.8 | --- | 4.0 | --- |
| male gender, n (%) | 793 (60.7) | 576 (60.0) | 141 (61.0) | 0.77 | 36 (69.2) | 0.19 |
| age, median (SD) | 67.2 (9.4) | 65.8 (9.0) | 71.2 (9.3) | <0.0001 | 72.2 (13.5) | <0.001 [a] |
| ECOG at Dx, mean (SD) | 0.85 (0.70) | 0.68 | 1.49 | <0.0001 | 1.13 | **<0.001 [a]** |
| 0, n (%) | 398 (30.5) | 360 (37.5) | 17 (7.4) | | 9 (17.3) | |
| 1, n (%) | 731 (56.0) | 554 (57.7) | 110 (47.6) | | 28 (53.8) | |
| 2, n (%) | 148 (11.3) | 43 (4.5) | 80 (34.6) | <0.0001 | 14 (26.9) | <0.0001 [a] |
| 3, n (%) | 27 (2.1) | 3 (0.3) | 22 (9.5) | | 1 (1.9) | |
| 4, n (%) | 2 (0.2) | 0 (0) | 2 (0.9) | | 0 (0) | |
| M1 sites at Dx, mean (SD) | 2.63 (2.0) | 2.5 (1.9) | 3.2 (2.2) | <0.0001 | 2.6 (1.9) | 0.74 |
| M1c cases, n (%) | 796 (60.9) | 575 (59.9) | 157 (68.0) | 0.024 | 33 (63.5) | 0.61 |
| Squamous, n (%) | 265 (20.3) | 194 (20.2) | 49 (21.2) | 0.73 | 10 (19.2) | 0.85 |
| Height cm, mean (SD) | 170.0 (9.1) | 171.4 (9.2) | 170.5 (8.9) | 0.18 | 172.0 (9.1) | 0.61 |
| Weight kg, mean (SD) | 72 (16.3) | 74.6 (16.5) | 70.4 (16.0) | 0.0006 | 73.3 (17.4) | 0.30 |
| BMI, mean (SD) | 24.4 (5.0) | 25.3 (5.1) | 24.1 (4.6) | 0.0005 | 24.7 (5.7) | 0.42 |
| Smoking status | | | | | | |
| never (n,%) | 112 (8.6) | 76 (7.9) | 23 (10.0) | | 5 (9.8) | |
| ex-smokers (n,%) | 690 (52.9) | 509 (53.0) | 126 (54.7) | 0.45 | 24 (47.1) | 0.69 |
| current (n,%) | 504 (38.5) | 375 (39.1) | 82 (35.3) | | 22 (43.1) | |
| Nr. of SOC, mean (SD) | 1.4 (1.2) | 1.31 (1.1) | 1.9 (1.3) | <0.0001 | 1.53 (1.2) | 0.094 |
| 0 (n,%) | 383 (29.3) | 318 (33.1) | 43 (18.6) | | 9 (17.3) | |
| 1 (n,%) | 319 (24.4) | 235 (24.5) | 46 (19.9) | <0.0001 | 21 (40.4) | 0.093 |
| 2 (n,%) | 329 (25.2) | 238 (24.8) | 64 (27.8) | | 11 (21.2) | |
| 3+ (n,%) | 277 (21.1) | 169 (17.6) | 78 (33.7) | | 11 (5.7) | |
| SCS, mean (SD) | 8.4 (3.1) | 8.2 (2.8) | 9.1 (3.8) | <0.0001 | 8.6 (3.5) | 0.28 |
| CCI, mean (SD) | 7.3 (1.3) | 7.2 (1.2) | 7.7 (1.5) | <0.0001 | 7.5 (1.1) | 0.15 |
| ACCI, mean (SD) | 9.6 (1.8) | 9.3 (1.7) | 10.3 (2.0) | <0.0001 | 10.0 (1.7) | 0.005 [a] |
| SmPC at Dx, n (%) | 501 (38.4) | 268 (27.9) | 162 (70.1) | <0.0001 | 30 (57.7) | <0.0001 [a] |
| SmPC before Tx/death, n (%) | 603 (46.2) | 304 (31.7) | 225 (97.4) | <0.0001 | 32 (61.5) | **<0.0001 [a]** |
| Δ(SmPC) before Tx/death - at Dx, n (%) | 116 (8.9) | 42 (4.4) | 69 (29.9) | <0.0001 | 4 (7.7) | 0.26 |
| Δ(ECOG) before Tx/death - at Dx, SD | 0.29 (0.45) | 0.24 (0.43) | 0.55 (0.50) | <0.0001 | 0.19 (0.40) | 0.42 |
| Δ(days) first CTT - first contact with hospital, SD | -11.7 (30.4) | -11,6 (33.6) | -11.7 (17.3) | 0.97 | -12.6 (25.1) | 0.83 |
| Δ(days) biopsy - first contact with hospital, SD | 6.8 (32.1) | 5.4 (34.4) | 10.1 (27.1) | 0.025 | 11.1 (17.4) | 0.037 [a] |
| Δ(days) Tx start or death - biopsy, SD | 49.7 (85.9) | 40.1 (45.4) | 52.9 (64.5) | 0.008 | 34.6 (25.0) | 0.050 |
| Δ(days) Tx start or death - biopsy, SD | 71.6 (135.9) | 52.9 (85.8) | 83.3 (102.4) | <0.0001 | 56.2 (41.1) | 0.30 |
| Radiotherapy before systemic therapy, n (%) | 329 (25.2) | 210 (21.9) | 80 (34.6) | <0.0001 | 22 (42.3) | 0.0007 [a] |
| NGS testing, n (%) [b] | 1056 (81.9) | 829 (86.4) | 142 (61.5) | <0.0001 | 34 (65.4) | <0.0001 [a] |
| NGS among nsq, % [c] | 89.4% | 94.9% | 69.8% | <0.0001 | 76.2% | <0.0001 [a] |
| mOS (95% CI) | 6.7 (5.9-7.4) | 10.6 (9.7-11.5) | 1.7 (1.5-1.9) | <0.0001 | 1.3 (1.0-1.7) | **<0.001 [a]** |

**Supplementary Table S2. Characteristics of the main study population according to therapeutic allocation (full version of Table 1)**

Significant differences ($p<0.05$) for the comparisons "mBSC" *vs.* ”any Tx", and "early death" *vs.* ”any Tx" are highlighted in italics; furthermore, for parameters with significant differences in both comparisons, those with significant difference also in the comparison "mBSC" *vs.* "early death" are highlighted in bold italics. Statistical comparisons were performed with a t-test for continuous and Chi-square test for categorical variables.

[a] for early death *vs.* mBSC: p=0.23 for age; p=0.002 for mean ECOG at Dx; p=0.058 for the ECOG at Dx distribution; p=0.12 for the ACCI; p=0.083 for the SmPC status at Dx, p<0.001 for the SmPC status before (planned) Tx or death; p=0.81 for the days between biopsy and first contact; p=0.30 for the initiation of RT before systemic therapy; p=0.60 for NGS testing; p=0.72 for NGS testing among non-squamous tumors; p=0.029 for OS.

[b] according to the reimbursement regulations, reflex testing is not allowed, *i.e.* NGS testing is intentionally triggered by the treating physicians after receipt of histological results whenever treatment is considered.

[c] NGS testing for squamous tumors was not generally recommended and widely practiced in Germany until a change in the national S3 guideline for lung cancer in 2022.

Abbreviations: ACCI: age-adjusted CCI; BMI: Body Mass Index; CCI: Charlson Comorbidity Index; CI: Confidence Interval; CTT: computerized tomography of the chest; Dx: diagnosis; ECOG PS: ECOG Performance Status; M1: distant metastasis; M1c: multiple extrathoracic metastases; mBSC: BSC due to medical reasons; mOS: median OS; n, N: number; NGS= Next-generation sequencing; NGS among nsq: NGS testing among non-squamous tumors; RT: radiation therapy, SCS: Simplified comorbidity score; SD: Standard Deviation; SOC System Organ Class affected by comorbidities; Tx: therapy; yr: years; Δ(): change in; ---: not applicable.

| | **ECOG PS before start of 1L-therapy or death** | | | | | |
|---|---|---|---|---|---|---|
| **ECOG PS at Dx** | **0** | **1** | **2** | **3** | **4** | **total** |
| **0** | 208 | 154 | 28 | 7 | 1 | 398 |
| **1** | 136 | 440 | 112 | 43 | 0 | 731 |
| **2** | 1 | 34 | 78 | 34 | 1 | 148 |
| **3** | 0 | 2 | 3 | 22 | 0 | 27 |
| **4** | 0 | 0 | 0 | 2 | 0 | 2 |
| **total** | 345 | 630 | 221 | 108 | 2 | 1306 |

#### Supplementary Table S3. ECOG PS at diagnosis vs. before start of 1L-therapy or death for total study cohort.

Number of patients shown per ECOG PS category. Pearson’s Chi-Square p <0.001.

Abbreviations: 1L: first-line; Dx: diagnosis; ECOG PS: ECOG Performance Status.

| **Reason for lack of platinum treatment in non-SmPC patients of the main study population (n=123 or 15.2%)** | **n (%)** | **% of all 1306** |
|---|---|---|
| mBSC due to clinical deterioration during workup | 80 (65) | 6.1 |
| early death before planned therapy | 15 (12) | 1.1 |
| BSC due to own wish | 23 (19) | 1.8 |
| young age <70 years with platinum contraindications [a] | 5 (4) | 0.4 |

#### Supplementary Table S4. Reasons for lack of platinum treatment in non-SmPC patients.

[a] e.g. chronic renal failure, severe coronary artery disease (CAD), and severe pulmonary disease (COPD GOLD IV).

Abbreviations: BSC: Best supportive care; mBSC: Best supportive care for medical reasons; n: number of patients; SmPC: Summary of medicinal Product Characteristics.

| Category | % all pts N=1306 | Non-SmPC at Dx n=805 | | SmPC at Dx n= 501 | |
|---|---|---|---|---|---|
| | | % of 805 (n) | mOS, 95% CI | % of 501 (n) | mOS, 95% CI |
| <u>Patient disposition</u> | | | | | |
| **All patients (n)** | 100 | 61.6 [a] (805) | 9.5 (8.4-10.6) | 38.4 [a] (501) | 4.2 (3.5-4.9) *** |
| **Any systemic Tx** | 73.5 | 86.0 (692) | 11.2 (9.9-12.5) | 53.5 (268) *** | 9.2 (7.7-10.7) ** |
| **Platinum-based Tx** | 70.4 | 84.7 (682) | 11.3 (10.1-12.5) | 47.5 (238) *** | 9.6 (7.9-11.3) * |
| **Non-platinum therapies** | 3.0 | 1.24 (10) | 4.5 (0.0-13.2) | 6.0 (30) *** | 5.1 (2.3-7.9) |
| **Early death** | 4.0 | 2.7 (22) | 1.4 (0.8-2.0) | 6.0 (30) ** | 1.3 (0.9-1.7) |
| **mBSC** | 17.7 | 8.6 (69) | 1.7 (1.5-2.0) | 32.3 (162) *** | 1.7 (1.4-2.0) |
| **BSC, patient's wish** | 4.6 | 2.7 (22) | 5.1 (1.9-8.3) | 7.6 (38) *** | 3.3 (1.9-4.8) |
| **NGS testing among all patients** | 80.9% (1056) | 85.1% (685) | 10.3 (9.0-11.6) | 74.1% (371/501) | 5.4 (4.6-6.2) *** |
| **NGS testing among treated patients** | 86.4% (829/960) | 87.4% (605/692) | 11.9 (10.6-13.2) | 83.6% (224/268) | 9.6 (8.0-11.2) ** |
| **NGS testing among non-treated patients** | 65.6% (227/346) | 70.8% (80/113) | 2.0 (1.9-2.1) | 63.1% (147/233) | 2.2 (1.9-2.5) |
| <u>SmPC categories</u> | | | | | |
| **>80 years old** | 129 (9.9) | 0 (0) | --- | 129 (25.7) *** | 4.5 (3.3-5.7) |
| **ECOG ≥3** | 29 (2.2) | 0 (0) | --- | 29 (5.8) *** | 1.5 (0.4-2.6) |
| **ECOG ≥2 + ≥1 SOC** | 116 (8.9) | 0 (0) | --- | 116 (23.2) *** | 1.8 (1.4-2.2) |
| **≥ 70 years + ≥1 SOC** | 421 (32.2) | 0 (0) | --- | 421 (84.0) *** | 5.0 (4.2-5.8) |
| **SOC 1+** | 923 (70.7) | 444 (55.1) | 9.3 (8.1-10.5) | 479 (95.6) *** | 4.2 (3.4-5.0) *** |
| **SOC 2+** | 604 (46.2) | 256 (31.8) | 8.0 (6.4-9.6) | 348 (69.3) *** | 3.8 (2.3-4.8) *** |
| **SOC 3+** | 275 (21.1) | 93 (11.6) | 6.7 (5.2-8.2) | 182 (36.3) *** | 3.1 (2.4-3.8) * |
| **Age ≥70** | 508 (38.9) | 66 (8.2) | 4.7 (1.6-7.8) | 442 (88.2) *** | 5.00 (4.3-5.7) |
| **ECOG ≥2** | 177 (13.6) | 30 (3.7) | 1.4 (1.1-1.7) | 147 (29.3) *** | 1.8 (1.5-2.1) |

**Supplementary Table S5. SmPC population: prevalence, characteristics and treatment.**

Statistically significant differences for the comparison "SmPC at Dx" *vs.* "non-SmPC at Dx" have been indicated with asterisks: *: p<0.05; **: p<0.01; ***: p<0.0001, all other p-values are >0.05. Statistical comparisons were performed with a Chi-square test for categorical variables and logrank test for survival.

[a] of all 1306 patients

<u>Abbreviations:</u> Best supportive care; CI: Confidence Interval; Dx: diagnosis; ECOG PS: ECOG Performance Status; IO: chemoimmunotherapy; mBSC: best supportive care for medical reasons; mOS: median overall survival; n, N:

number; NGS: Next-generation sequencing; pts: patients; SmPC: Summary of medicinal Product Characteristics; SOC: System Organ Class, number of systems affected by comorbidities; Tx: treatment; ---: not applicable.

| Platinum treatment | % all pts | non-SmPC pts | mOS, 95% CI | SmPC pts | mOS, 95% CI |
|---|---|---|---|---|---|
| **combined with IO, n (%)** [a] | 686 (74.6) | 504 (73.9) | 12.8 (11.3-14.3) | 182 (76.5) | 10.2 (8.0-12.4) ** |
| **cycles nr, mean (SE)** | 3.1 (0.04) | 3.1 (0.05) | --- | 2.8 (0.08) *** | --- |
| **dose ratio for cycles 1-4, mean % (SE)** | 91.1 (0.03) | 93.4 (0.003) | --- | 84.3 (0.007) *** | --- |
| **DR at c1, % (n)** | 32.7 (301) | 22.3 (152) | 12.8 (11.6-14.0) | 62.6 (149) *** | 7.8 (6.4-9.2) *** |
| **DR under Tx, % (n)** | 19.6 (180) | 20.4 (139) | 13.2 (11.2-15.2) | 17.2 (41) | 10.2 (9.0-11.4) * |
| **DR anytime, % (n)** | 49.6 (456) | 40.8 (278) | 11.9 (10.1-13.7) | 74.8 (178) *** | 10.2 (8.9-11.5) ** |
| **DI, % pts** | 27.3 (248) | 25.2 (170) | 11.7 (10.5-12.9) | 33.3 (78) * | 9.3 (8.0-10.6) * |
| **DR or DI, % (n)** | 60 (552) | 52.3 (357) | 13.1 (11.3-15.0) | 81.9 (195) *** | 9.8 (8.6-11.0) *** |
| **DR & DI, % (n)** | 16.5 (152) | 13.3 (91) | 11.2 (10.1-12.3) | 25.6 (61) *** | 9.9 (8.0-11.8) |
| **DR or DI or DC, % (n)** | 72.6 (668) | 66.4 (453) | 19.0 (14.3-23.7) | 90.3 (215) *** | 8.3 (7.3-9.3) *** |
| **DR & DI & DC, % (n)** | 8.6 (79) | 6.7 (46) | 11.2 (10.1-12.3) | 13.9 (33) *** | 6.8 (5.3-8.3) ** |
| **Grade 3 AE, %** | 34.3 (316) | 33.0 (225) | 10.7 (9.4-12.0) | 38.2 (91) | 11.1 (9.4-12.8) |

**Supplementary Table S6. SmPC population: platinum exposure and toxicity.**

Statistically significant differences have been indicated with asterisks: *: p<0.05; **: p<0.01; ***: p<0.0001 for SmPC patients vs. non-SmPC patients, all other p-values are >0.05. Statistical comparisons were performed with a logrank test for survival.

[a] some patients received platinum doublets alone before approval of chemoimmunotherapy by the EMA, due to contraindications for immune checkpoint inhibitors *etc.*

Abbreviations: AE: Adverse event; c1: first cycle; CI: Confidence Interval; DC: platinum discontinuation; DI: dose interruption; DR: dose reduction; Dx: diagnosis; ECOG PS: ECOG Performance Status; mOS: median overall survival; n, N: number; pts: patients; SE: Standard error; SmPC: Summary of medicinal Product Characteristics; Tx: treatment; ---: not applicable.

| | SmPC patients with average AUC ≤3 (n=119) | SmPC patients with average AUC >3 (n=119) | Platinum-treated non-SmPC patients (n=682) | p-value |
|---|---|---|---|---|
| **DR at c1, %** | 73.9% | 51.3% | 22.3% | p<0.001 |
| **DR or DI or DC, %** | 98.3% | 82.4% | 66.4% | p<0.001 |
| **DR and DI and DC, %** | 24.4% | 3.4% | 6.7% | p<0.001 |
| **DC due to toxicity with subsequent other therapy** | 45.4% | 7.6% | 8.4% | p<0.0001 |
| **DC with subsequent death (w/o radiologic progression)** | 26.9% | 0.8% | 10.7% | p<0.0001 |
| **DC with radiologic tumor progression** | 13.4% | 3.4% | 17.2% | p=0.0004 |
| **mPFS, months** | 3.7 (3.0 – 4.4) | 8.0 (6.7 – 9.3) | 5.6 (5.1 – 6.1) | p<0.001 |
| **mOS, months** | 5.1 (3.9 – 6.3) | 15.5 (11.5 – 19.5) | 11.3 (10.1 – 12.5) | p<0.001 |

**Supplementary Table S7. Platinum dose adjustments, discontinuation and survival for platinum-treated SmPC patients with low average carboplatinum dose AUC ≤3 *vs.* higher platinum dose vs. platinum-treated non-SmPC patients.**

Dosing details are given in the Suppl. Figure S2. Statistical comparisons were performed with a Chi-square test for categorical data and logrank test for survival.

Abbreviations: AUC: area-under-the-curve; c1: first cycle; DC: platinum discontinuation; DI: dose interruption; DR: dose reduction; mPFS: median progression-free survival; mOS: median overall survival; n: number; SmPC: Summary of medicinal Product Characteristics

| | Non-SmPC at Dx N=805 | | | SmPC at Dx N= 501 | | |
|---|---|---|---|---|---|---|
| | **% of 805** | **mPFS, 95% CI** | **mOS, 95% CI** | **% of 501** | **mPFS, 95% CI** | **mOS, 95% CI** |
| **Pemetrexed** | 0.1 (1) | 4.5 (---) [a] | 4.5 (---) [a] | 1.6 (8) | 1.5 (0.8-2.2) | 1.5 (0.02-30) |
| **Gemcitabine** | 0.7 (6) | 3.6 (0-8.5) | 6.1 (0-15.1) | 2.4 (12) | 4.5 (3.3-5.7) | 9.0 (3.1-14.9) |
| **Vinorelbine** | 0.1 (1) | 10.8 (---) [a] | 10.8 (---) [a] | 1.0 (5) | 1.8 (1.6-2.0) | 3.0 (2.6-3.4) |
| **Vincristine-Etoposide** | 0.1 (1) | 0.5 (---) [a] | 0.5 (---) [a] | 0.4 (2) | 3.6; 5.1(---) [a] | 5.1; 13.7 (---) [a] |
| **Anti PD-(L)1 antibody** | 0.1 (1) | 0.5 (---) [a] | 0.5 (---) [a] | 0.6 (3) | 2.7 (0-6.4) | 3. 0 (0.0-6.2) |

**Supplementary Table S8. Details of non-platinum therapies in study patients (n=40).**

[a] due to the small sample size (n<3), individual values are listed (all with OS event)

Abbreviations: CI: Confidence Interval; Dx: diagnosis; mOS: median overall survival; mPFS: median progression-free survival; N: number; OS: overall survival; PD-(L)1: Programmed Death-Ligand 1; SmPC: Summary of medicinal Product Characteristics; ---: not applicable

| | Patient subset without platinum treatment to be predicted<br>All patients<br>n=386 | <br>All patients<br>with NGS n=258 | <br>Nonsquamous<br>with NGS n=230 |
|---|---|---|---|
| **reference** | all patients<br>(N=1306) | patients with NGS<br>(N=1056) | non-squamous with NGS<br>(N=931) |
| | **AUC (95% CI)<br>p-value; Gini index** | **AUC (95% CI)<br>p-value; Gini index** | **AUC (95% CI)<br>p-value; Gini index** |
| ***SmPC criteria at Dx*** | *0.708 (0.675-0.741)<br>p<0.0001; 0.416* | *0.701 (0.662-0.739)<br>p<0.0001; 0.401* | *0.695 (0.654-0.736)<br>p<0.0001; 0.390* |
| **ECOG PS ≥2 at Dx** | 0.662 (0.625-0.699)<br>p<0.0001; 0.324 | 0.647 (0.604-0.690)<br>p<0.0001; 0.293 | 0.637 (0.592-0.683)<br>p<0.0001; 0.275 |
| **Age ≥70 at Dx** | 0.657 (0.623-0.691)<br>p<0.0001; 0.314 | 0.666 (0.627-0.705)<br>p<0.0001; 0.331 | 0.662 (0.620-0.704)<br>p<0.0001; 0.324 |
| **SCS** | 0.601 (0.564-0.638)<br>p<0.0001; 0.202 | 0.589 (0.546-0.632)<br>p<0.0001; 0.177 | 0.583 (0.537-0.628)<br>p<0.0001; 0.165 |
| **CCI** | 0.597 (0.562-0.632)<br>p<0.0001; 0.193 | 0.575 (0.534-0.615)<br>p<0.0001; 0.150 | 0.590 (0.547-0.633)<br>p=0.001; 0.180 |
| **ACCI** | 0.673 (0.641-0.706)<br>p<0.0001; 0.346 | 0.663 (0.626-0.701)<br>p<0.0001; 0.326 | 0.673 (0.633-0.712)<br>p<0.0001; 0.345 |
| **BMI** | 0.443 (0.408-0.478)<br>p=0.001; -0.113 | 0.460 (0.419-0.500)<br>p=0.052; -0.080 | 0.460 (0.417-0.503)<br>p=0.067; -0.080 |
| **SOC, nr.** | 0.624 (0.590-0.659)<br>p<0.0001; 0.248 | 0.614 (0.574-0.654)<br>p<0.0001; 0.228 | 0.624 (0.582-0.667)<br>p<0.0001; 0.249 |
| **Serum creatinine at Dx** | 0.543 (0.505-0.580)<br>p=0.026; 0.085 | 0.552 (0.508-0.596)<br>p=0.019; 0.104 | 0.541 (0.495-0.588)<br>p=0.083; 0.083 |
| **eGFR by MDRD at Dx** | 0.441 (0.403-0.478)<br>p=0.002; -0.118 | 0.423 (0.380-0.467)<br>p=0.001; -0.153 | 0.438 (0.391-0.485)<br>p=0.009; -0.124 |
| **Metastatic sites, nr.** | 0.553 (0.518-0.589)<br>p=0.005; 0.107 | 0.560 (0.518-0.601)<br>p=0.005; 0.119 | 0.573 (0.529-0.616)<br>p=0.0.001; 0.146 |

**Supplementary Table S9. The predictive performance of SmPC criteria and various clinical characteristics for the identification of patients who will not receive platinum.**

SmPC criteria showed the parameters with the best performance and have been highlighted in italics. The sensitivity and specificity of SmPC criteria for the three predictions from left to right were 68.1% and 74.1%, 65.9% and 74.8%; 63.5% and 76.2%, respectively.

Abbreviations: ACCI: age-Adjusted CCI; AUC: area-under-the-curve; BMI: Body mass index; CCI: Charlson Comorbidity Index ; Dx: diagnosis; ECOG PS: ECOG Performance Status; eGFR: epidermal Growth Factor Receptor; MDRD: Modification of Diet in Renal Disease formula; n, N: number; NGS: Next-generation sequencing; nr: number; SCS: Simplified Comorbidity Score; SmPC: Summary of medicinal Product Characteristics; SOC: System Organ Class.

| Patient subset to be predicted | Platinum-treated patients with platinum DC due to toxicity (n=120) or death (n=106) | Platinum-treated SmPC patients with platinum DC due to toxicity (n=63) or death (n=33) |
|---|---|---|
| **reference** | all platinum-treated patients (N=920) | platinum-treated SmPC patients (N=238) |
| | **AUC (95% CI)<br>p-value; Gini index** | **AUC (95% CI)<br>p-value; Gini index** |
| **SmPC criteria at Dx** | 0.613 (0.568-0.657)<br>p<0.001; 0.225 | n/a |
| **SmPC criteria before Tx/death** | 0.621 (0.568-0.657)<br>p<0.001; 0.242 | n/a |
| **ECOG PS ≥2 at Dx** | 0.538 (0.492-0.583)<br>p=0.103; 0.075 | 0.547 (0.471-0.624)<br>p=0.224; 0.095 |
| **DR in the first cycle** | 0.628 (0.584-0.671)<br>p<0.001; 0.255 | 0.553 (0.478-0.628)<br>p=0.164; 0.107 |
| **Age ≥70 at Dx** | 0.601 (0.557-0.646)<br>p<0.001; 0.202 | 0.469 (0.393-0.546)<br>p=0.433; -0.061 |
| **SCS** | 0.537 (0.492-0.581)<br>p=0.106; 0.074 | 0.559 (0.484-0.635)<br>p=0.123; 0.119 |
| **CCI** | 0.571 (0.528-0.615)<br>p=0.001; 0.143 | 0.475 (0.399-0.552)<br>p=0.525; -0.050 |
| **ACCI** | 0.626 (0.585-0.667)<br>p<0.001; 0.253 | 0.468 (0.391-0.546)<br>p=0.421; -0.063 |
| **BMI** | 0.496 (0.452-0.540)<br>p=0.851; -0.008 | 0.461 (0.385-0.537)<br>p=0.311; -0.079 |
| **SOC, n** | 0.570 (0.527-0.612)<br>p=0.001; 0.139 | 0.502 (0.426-0.577)<br>p=0.965; 0.003 |
| **Serum creatinine at Dx** | 0.555 (0.509-0.601)<br>p=0.019; 0.110 | 0.559 (0.483-0.636)<br>p=0.128; 0.119 |
| **eGFR by MDRD at Dx** | 0.413 (0.367-0.458)<br>p<0.001; -0.175 | 0.415 (0.340-0.491)<br>p=0.027; -0.169 |
| **Metastatic sites, nr.** | 0.560 (0.516-0.603)<br>p=0.007; 0.120 | 0.527 (0.452-0.602)<br>p=0.485; 0.053 |

**Supplementary Table S10. The predictive performance SmPC criteria and various clinical characteristics for the identification of platinum-treated patients with discontinuation due to toxicity or death before completing 4 chemotherapy cycles.**

The sensitivity and specificity of SmPC criteria at Dx or before Tx were 42.5% and 79.5%, or 46.9% and 76.74%, respectively..

Abbreviations: ACCI: age-Adjusted CCI; BMI: Body mass index; CCI: Charlson Comorbidity Index ; DC: discontinuation; DR: dose reduction; Dx: diagnosis; ECOG PS: ECOG Performance Status; eGFR: epidermal Growth Factor Receptor; MDRD: Modification of Diet in Renal Disease formula; n, N: number; SCS: Simplified Comorbidity Score; SmPC: Summary of medicinal Product Characteristics; SOC: System Organ Class affected by comorbidities; Tx: therapy.

| Patient subset to be predicted | SmPC starting with AUC4 and receiving <4 cycles, n=76 | SmPC starting with AUC5 and receiving 1-2 cycles, n=31 | All SmPC patients with mAUC≤3 n=119 |
|---|---|---|---|
| reference | all SmPC starting with AUC4 (N=136) | all SmPC starting with AUC5 (N=89) | SmPC with platinum (N=238) |
| | **AUC (95% CI) p-value; Gini index** | **AUC (95% CI) p-value; Gini index** | **AUC (95% CI) p-value; Gini index** |
| **ECOG PS ≥2 at Dx** | 0.567 (0.469-0.688) p=0.181; 0.132 | 0.539 (0.408-0.671) p=0.556; 0.079 | 0.550 (0.476-0.625) p=0.182; 0.101 |
| **DR in the first cycle** | 0.500 (0.401-0.599) p=1.00; 0.00 | n/a | 0.613 (0.540-0.686) p=0.002; 0.226 |
| **Age ≥70 at Dx** | 0.448 (0.623-0.691) p=0.299; -0.103 | 0.477 (0.347-0.608) p=0.733; -0.045 | 0.466 (0.392-0.541) p=0.374; -0.067 |
| **SCS** | 0.544 (0.445-0.644) p=0.382; 0.089 | 0.492 (0.359-0.625) p=0.904; -0.016 | 0.525 (0.451-0.600) p=0.507; 0.050 |
| **CCI** | 0.462 (0.364-0.561) p=0.452; -0.075 | 0.419 (0.289-0.550) p=0.227; -0.161 | 0.460 (0.385-0.534) p=0.289; -0.080 |
| **ACCI** | 0.446 (0.349-0.543) p=0.277; -0.108 | 0.448 (0.315-0.581) p=0.443; -0.104 | 0.469 (0.394-0.544) p=0.416; -0.062 |
| **BMI** | 0.484 (0.385-0.582) p=0.743; -0.033 | 0.522 (0.390-0.654) p=0.746; 0.044 | 0.484 (0.409-0.558) p=0.668; -0.033 |
| **SOC, nr.** | 0.496 (0.397-0.596) p=0.941; -0.007 | 0.468 (0.337-0.600) p=0.638; -0.063 | 0.496 (0.422-0.571) p=0.924; -0.007 |
| **Serum creatinine at Dx** | 0.531 (0.432-0.629) p=0.546; 0.061 | 0.578 (0.445-0.710) p=0.252; 0.155 | 0.553 (0.478-0.627) p=0.164; 0.106 |
| **eGFR by MDRD at Dx** | 0.449 (0.351-0.547) p=0.310; -0.101 | 0.407 (0.277-0.537) p=0.160; -0.186 | 0.422 (0.349-0.496) p=0.039; -0.155 |
| **Metastatic sites, nr.** | 0.593 (0.497-0.689) p=0.058; -0.103 | 0.566 (0.439-0.694) p=0.309; 0.133 | 0.567 (0.493-0.641) p=0.074; 0.134 |

**Supplementary Table S11. The predictive performance of various characteristics and scores for the identification of patients with potential platinum overtreatment.**

Abbreviations: ACCI: age-Adjusted CCI; AUC: area-under-the-curve; BMI: Body mass index; CCI: Charlson Comorbidity Index ; Dx: diagnosis; ECOG PS: ECOG Performance Status; eGFR: epidermal Growth Factor Receptor; mAUC: mean AUC dose in the first 4 cycles; MDRD: Modification of Diet in Renal Disease formula; n, N: number; nr: number; SCS: Simplified Comorbidity Score; SmPC: Summary of medicinal Product Characteristics; SOC: System Organ Class

### Supplementary Figures

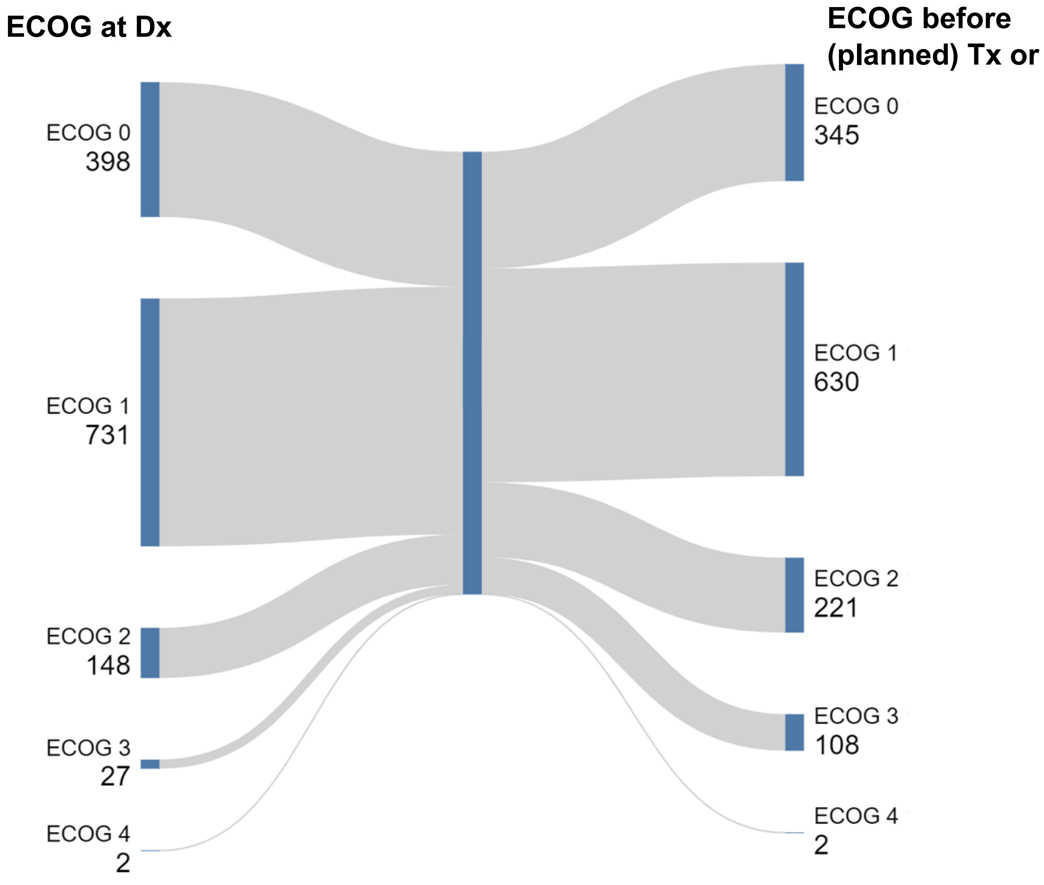


**Supplementary Figure S1: Changes in ECOG PS from initial contact until before start of (planned) treatment or death.**

Sankey plot showing changes in the ECOG Performance Status for patients with NSCLC PD-L1 TPS 0-49 without targeted options between initial diagnosis and time of planned treatment. Further details are given in Supplementary Table S3.

Abbreviations: ECOG 1L: ECOG Performance Status before start of first-line therapy or death; ECOG at Dx: ECOG Performance Status at the time of initial diagnosis; NSCLC: non-small-cell lung cancer; PD-L1: Programmed Death-Ligand 1; TPS: Tumor Proportion Score.

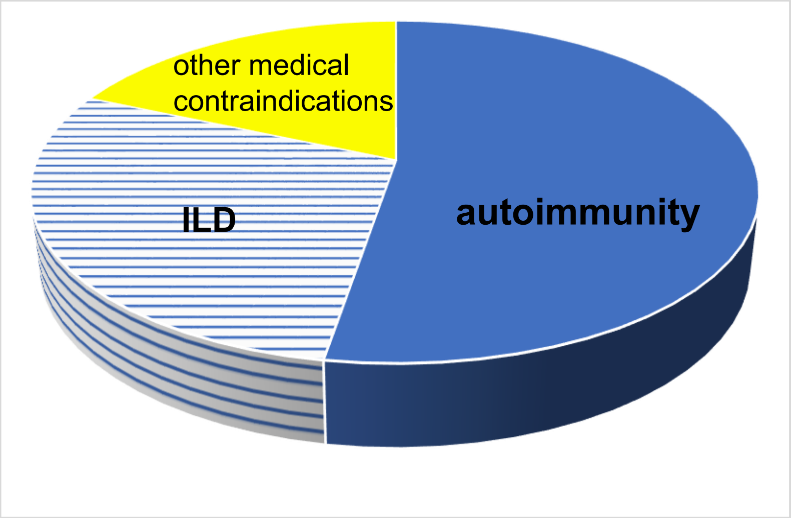


**Supplementary Figure S2: Contraindications to immunotherapy in NSCLC PD-L1 TPS 0-49 patients in this study**.

Immunotherapy contraindications were present in 4.7% (38/801) patients (NB. patients treated with platinum before immunotherapy approval for the specific histology or in a setting without availability of immunotherapy, e.g. within a clinical trial or without evidence of disease after definitive local therapy, were excluded from this analysis). Most prevalent cause was autoimmune disease (n=20), followed by interstitial lung disease (n=11) and other medical contraindications to immunotherapy (n=7, in particular poor respiratory function)

Abbreviations: ILD: Interstitial lung disease; NSCLC: Non-Small Cell Lung Cancer; PD-L1: Programmed Death-Ligand 1; TPS: Tumor Proportion Score.

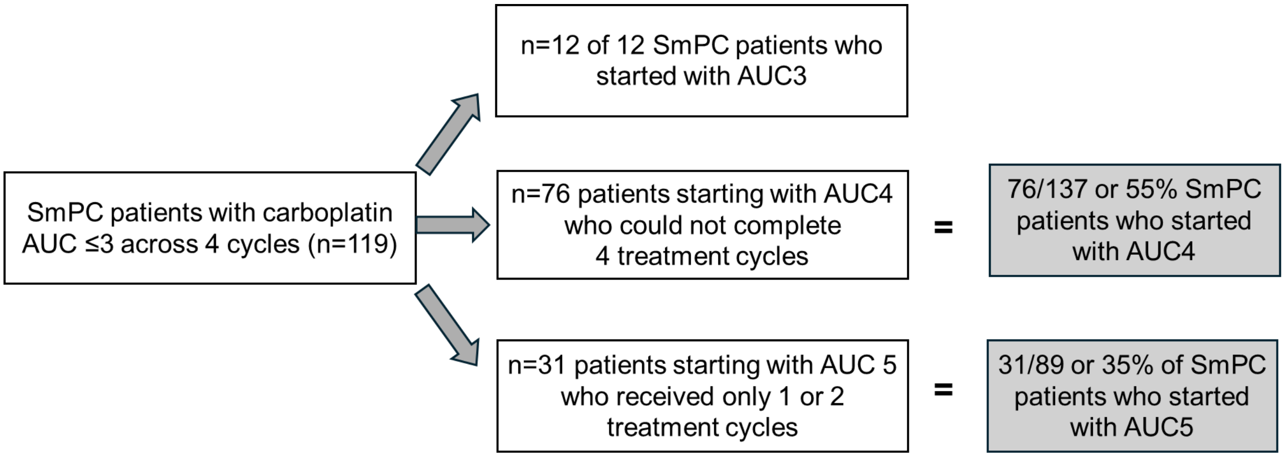


**Supplementary Figure S3. Composition of the SmPC patient group with low platinum dose (average AUC≤3 across 4 cycles).**

Abbreviations: AUC: area-under-the-curve, SmPC: Summary of medicinal Product Characteristics.

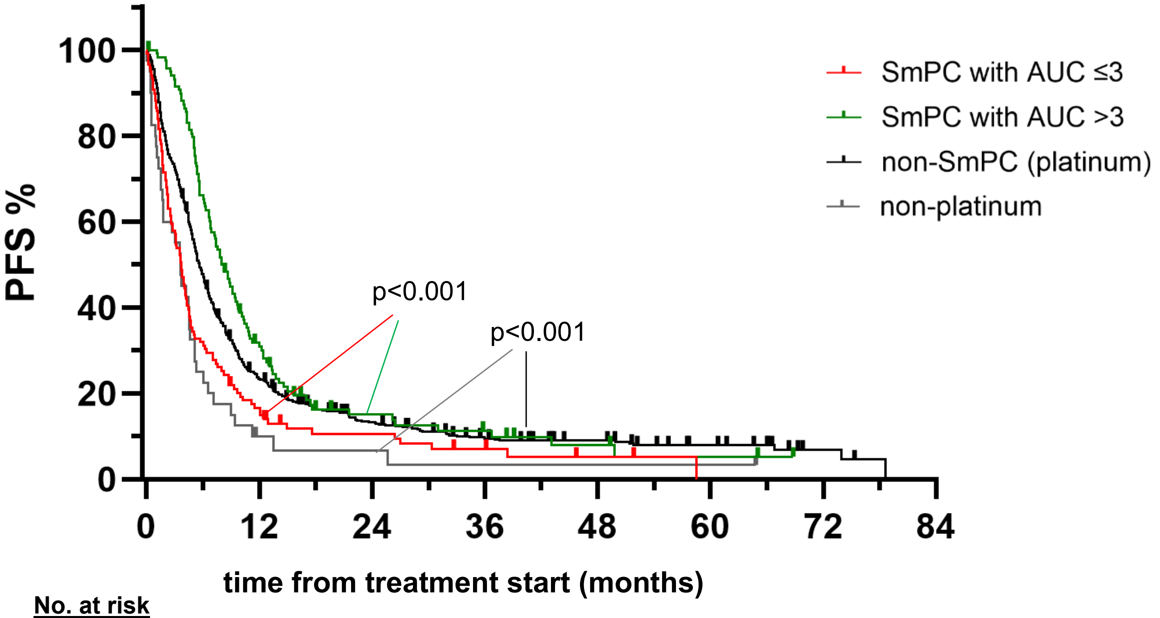


**No. at risk**

| | | | | | | | | |
|---|---|---|---|---|---|---|---|---|
| SmPC AUC ≤3: | 119 | 19 | 9 | 5 | 2 | 0 | | |
| SmPC AUC >3: | 119 | 36 | 12 | 8 | 4 | 2 | 0 | |
| Non-SmPC platinum: | 682 | 153 | 68 | 41 | 28 | 17 | 3 | 0 |
| Non-platinum: | 40 | 3 | 2 | 1 | 1 | 1 | 0 | |

**Supplementary Figure S4: Progression-free survival according to treatment.**

Median PFS was 3.7 (95% CI 3.0 – 4.4) months for SmPC patients with average AUC ≤3 (n=119), 8.0 (6.7 – 9.3) months for SmPC patients with average AUC >3 (n=119), 5.8 (5.1 – 8.1) months for platinum-treated non-SmPC patients (n=682) and 3.6 (2.5 – 4.7) for patients who received monotherapies (logrank $p<0.001$).

Abbreviations: CI: confidence interval; HR: hazard ratio; PFS: progression-free survival; SmPC: Summary of Product Characteristics for atezolizumab; Tx: therapy.
